# Systematic review and meta-analysis to estimate the burden of non-fatal and fatal overdose among people who inject drugs living in the U.S. and comparator countries: 2010 – 2023

**DOI:** 10.1101/2024.08.14.24310813

**Authors:** Jalissa Y. Shealey, Eric W. Hall, Therese D. Pigott, Lexi Rosmarin, Anastasia Carter, Chiquita Cade, Nicole Luisi, Heather Bradley

## Abstract

**Background:** People who inject drugs (PWID) have high risk for overdose, but there are no current estimates of overdose rates in this population. We estimated the rates of non-fatal and fatal overdose among PWID living in the U.S. and comparator countries (Canada, Mexico, United Kingdom, Australia), and ratios of non-fatal to fatal overdose, using literature published 01/01/2010 – 09/29/2023.

**Methods:** PubMed, PsychInfo, Embase, and ProQuest databases were systematically searched to identify publications reporting prevalence or rates of recent (past 12 months) non-fatal and fatal overdose among PWID. Non-fatal and fatal overdose rates were meta-analyzed using random effects models. Risk of bias was assessed using an adapted quality assessment tool, and heterogeneity was explored using sensitivity analyses.

**Results:** Our review included 143 records, with 58 contributing unique data to the meta-analysis. Non-fatal and fatal overdose rates among PWID in the U.S. were 32.9 per 100 person-years (PY) (95% CI: 26.4 – 40.9; n=28) and 1.7 per 100 PY (95% CI: 0.9 – 3.2; n=4), respectively. Limiting the analysis to data collected after 2016 yielded a non-fatal rate of 41.0 per 100 PY (95% CI: 32.1 – 52.5; n=16) and a fatal rate of 2.5 per 100 PY (95% CI: 1.4 – 4.3; n=2) in the U.S. An estimated 5% of overdoses among PWID in the U.S. result in death. Among the analyzed countries, Australia had the lowest non-fatal and fatal overdose rates and the largest ratio of non-fatal to fatal overdose.

**Conclusion:** Findings demonstrate substantial burden of non-fatal and fatal overdose among PWID in the U.S. and comparator countries. Scale-up of interventions that prevent overdose mortality and investments in PWID health research are urgently needed.

## Introduction

People who inject drugs (PWID) have high risk for overdose, leading to adverse health outcomes including preventable death. It is estimated that, globally, 18.5% of PWID experience at least one non-fatal overdose over the course of a year (Degenhardt et al., 2023). Compared to people who use drugs (PWUD) via other consumption routes, PWID are more likely to experience both non-fatal and fatal overdose due to rapid onset of effects (Caudarella et al., 2016; Mars et al., 2014). Physical consequences of non-fatal overdose can be severe (e.g., cognitive impairment, peripheral neuropathy, paralysis of limbs) and result in lifelong morbidity (Park et al., 2020; Warner-Smith et al., 2002). Consequently, healthcare utilization costs of overdose are substantial. In 2017, the economic burden of overdose and opioid use disorder (OUD) in the United States (U.S.) was estimated at $1.02 trillion, with more than 85% of costs attributable to the value of reduced quality of life and loss of life (Florence et al., 2021). Drug overdose deaths increased substantially during the COVID-19 pandemic (CDC, 2020), which led to an estimated 30% increase in the economic burden of fatal overdose from 2019 – 2020 (Lui et al., 2022). Non-fatal overdose events tend to predict fatal overdose (Caudarella et al., 2016). While burden of overdose among PWUD is estimable using surveillance data, there are limited current data on the burden of non-fatal and fatal overdose among PWID in the U.S.

Injection drug use (IDU) increased in the U.S. during the past decade due to the trajectory of the opioid epidemic. While the first U.S. wave (1990-2009) of the opioid epidemic involved primarily prescription opioids, the second (2010-2012) and third (2013-) waves are characterized to a large extent by use of commonly injected substances (e.g., heroin, synthetic opioids including fentanyl) (CDC, 2023c). In the U.S., it is estimated that 3.7 million people injected drugs in 2018, a 3 – 5 -fold increase from 2011 (Bradley et al., 2023), and it is still unclear how IDU prevalence may have further changed during the COVID-19 pandemic. There is an urgent need to understand how non-fatal and fatal overdose prevalence has changed among PWID alongside growth in this population.

Overdoses among PWID can be prevented with evidence-based interventions. Harm reduction interventions such as syringe services programs (SSPs) and naloxone distribution, as well as medication for opioid use disorder (MOUD) when appropriate, reduces risk for overdose and other adverse health outcomes among PWID (Adams et al., 2019; Buresh et al., 2020; Strange et al., 2022). Canada, the United Kingdom (U.K.), and Australia, have sanctioned operation of supervised consumption sites (SCSs) which may reduce overdose mortality and increase substance use treatment uptake (Kennedy, Hayashi, et al., 2019). A better understanding of the burden of overdose among PWID in the U.S. is critical for informing resource allocation to harm reduction and other overdose prevention interventions.

In 2019, the U.S. Centers for Disease Control and Prevention (CDC) launched the Overdose Data to Action (OD2A) initiative to improve the accuracy, comprehensiveness, and timeliness of overdose data in the U.S. (CDC, 2024d). Investment in these systems has improved the quality of overdose reporting, however, surveillance systems are currently unable to attribute overdoses to injection versus other consumption routes (Casillas et al., 2024).

National HIV Behavioral Surveillance (NHBS) is currently the only source of surveillance data on non-fatal overdose among PWID, but NHBS is conducted among PWID only every three years and in selected urban areas. Meta-analyses using research study data can be used to estimate the burden of overdose among PWID. For example, Degenhardt et al. (2023) conducted a global systematic review of non-fatal overdose and included studies published before 2010 for the U.S. estimate due to the prioritization of data coverage over recency (Colledge et al., 2019). Other meta-analyses estimated overdose among people with OUD regardless of route of administration (Bahji et al., 2020) or focused more broadly on all-cause mortality among PWID without providing country-level estimates (Larney et al., 2020; Mathers et al., 2013).

We expanded the scope of previously published meta-analyses (Bahji et al., 2020; Degenhardt et al., 2023; Larney et al., 2020; Mathers et al., 2013) by estimating the burden of non-fatal and fatal overdose in the U.S. and comparator countries (Canada, Mexico, United Kingdom, Australia) and using literature published between 2010 to 2023. In this systematic review, we used the same time period and meta-analytic methods to compute the ratio of non-fatal to fatal overdose among PWID, which can elucidate the proportion of overdoses that result in death and guide interventional strategy (Casillas et al., 2024). This systematic review and meta-analysis is aimed to:

1. Estimate rates of non-fatal and fatal overdose among PWID in the U.S. and comparator countries using data from studies published during 2010 – 2023.
2. Estimate time-specific rates of non-fatal and fatal overdose among PWID in the U.S. and comparator countries using data collected after 2016.
3. Estimate the ratio of non-fatal to fatal overdose among PWID in the U.S. and comparator countries, overall and post-2016.

## Methods

This systematic review and meta-analysis was conducted following the 2020 Preferred Reporting Items for Systematic Reviews and Meta-Analyses (PRISMA) statement guidelines (Page et al., 2021). The completed PRISMA checklist can be found in Appendix 1 of the supplemental material.

### Search Strategy

MEDLINE via PubMed, PsychInfo via EBSCO, Embase, and ProQuest databases were systematically searched to identify articles published from January 1, 2010 through September 29, 2023. Search strings included key terms related to drug injection (e.g., “PWID”, “IDU”, “injection”) and overdose or death (e.g., “mortality”, “fatal”). Relevant inclusion and exclusion terms were incorporated into our search strings using the search strategies of previously published meta-analyses as models (Bahji et al., 2020; Larney et al., 2020). These strings were tailored to each database searched in consultation with a systematic review librarian (Appendix 2). We also examined the reference list of a related review estimating the global prevalence of non-fatal overdose among PWID to identify citations not captured in our search (Degenhardt et al., 2023).

### Study Selection Criteria

Search results were imported into *Covidence Systematic Review Software* (2023), a web-based collaboration software platform that streamlines the production of systematic and other literature reviews, where duplicate citations were removed. Citations were excluded if [1] they were not published in English language, [2] study participants were not human, [3] data were collected exclusively from persons aged <18 years, [4] data were collected exclusively before 2010, [5] data were not collected in North America, the UK, or Australia, [6] there was no prevalence or rates of non-fatal or fatal overdose exclusive to people who inject drugs non-medically, [7] only included case reports or numerators for overdoses without population denominators, [8] only included lifetime (e.g., “ever”) PWID and did not report outcomes among recent (past 12 months) PWID, [9] only included lifetime (e.g., “ever”) non-fatal overdose prevalence or rates and did not report recent (past 12 months) non-fatal overdose, [10] findings included unconfirmed fatal overdose events, and [11] recall period for non-fatal overdose was less than 3 months or unknown. We limited our review to North America, the UK, and Australia, because we were interested in estimates of non-fatal and fatal overdose rates that could be compared to the U.S. Our previous review on this topic indicated these geographic areas were likely to have sufficient data for fatal and non-fatal overdose estimated (Shealey et al., 2022).

Using these exclusion criteria, titles and abstracts, followed by full-text articles were screened for eligibility by two independent reviewers (JYS, HB, EH, MG, AC, CC, and LR). Conflicts were resolved collaboratively through discussion or by a third reviewer.

### Data Extraction

Data from all eligible citations were extracted and coded by two independent reviewers using *Microsoft Excel* (2024). Conflicts were resolved collaboratively or by a third reviewer. The following data elements were extracted: the number of current PWID experiencing a non-fatal or fatal overdose; total years of follow-up; recall periods for both IDU and non-fatal overdose; sample size; overdose measurement method; definition used to describe non-fatal overdose; cohort or study name for studies conducted among established cohorts or studies; participant recruitment site(s); geographic location; data collection year(s); publication year; and first author name.

In cases where multiple citations presented data from the same cohort, the study with the most recent (i.e., data collection dates) and complete (i.e., sample size) data was included in the meta-analysis. When both baseline and follow-up data following an intervention to reduce overdose risk were reported, the baseline estimate was used. Decisions on the studies included in the meta-analysis were made by two independent reviewers. Conflicts were resolved through discussion or by a third reviewer.

### Quality Assessment

The adapted Newcastle-Ottawa Scale was used to assess risk of bias in non-randomized studies (Wells et al., 2013). We evaluated the quality of studies by rating them as high, low, or moderate risk of bias based on the following criteria: [1] participant recruitment method (population-based versus venue-based), [2] sample size (≥200 participants versus ≤199 participants), [3] whether person-years (PY) were reported directly or estimated by study team (for non-fatal overdose only), and [4] the clarity of the survey instrument’s definition of an “overdose” event (for non-fatal overdose only) (Figure 2). Two reviewers assessed study quality independently, resolving conflicts with a third reviewer.

**Figure 1.**
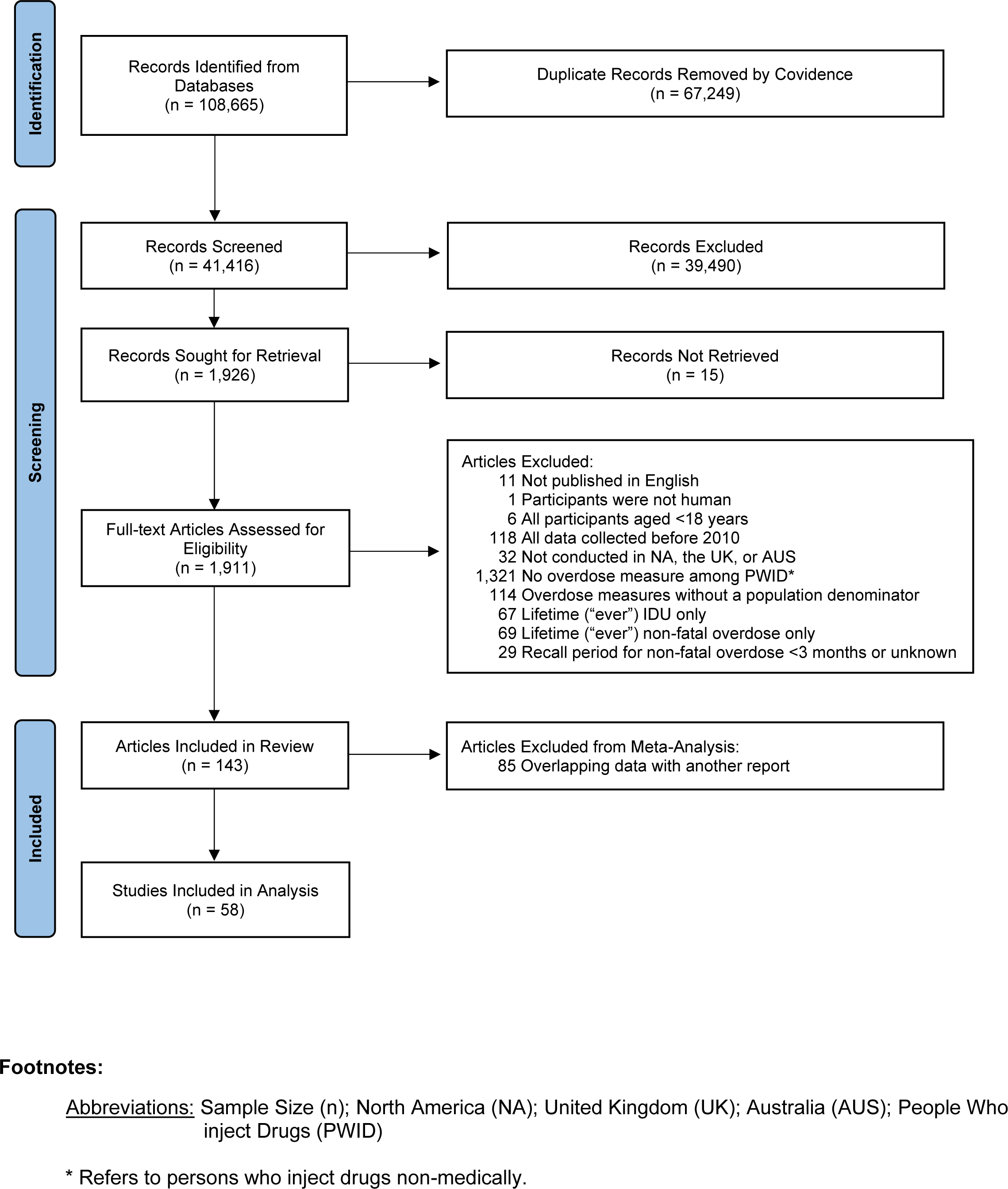
PRISMA Flow Chart

**Figure 2.**
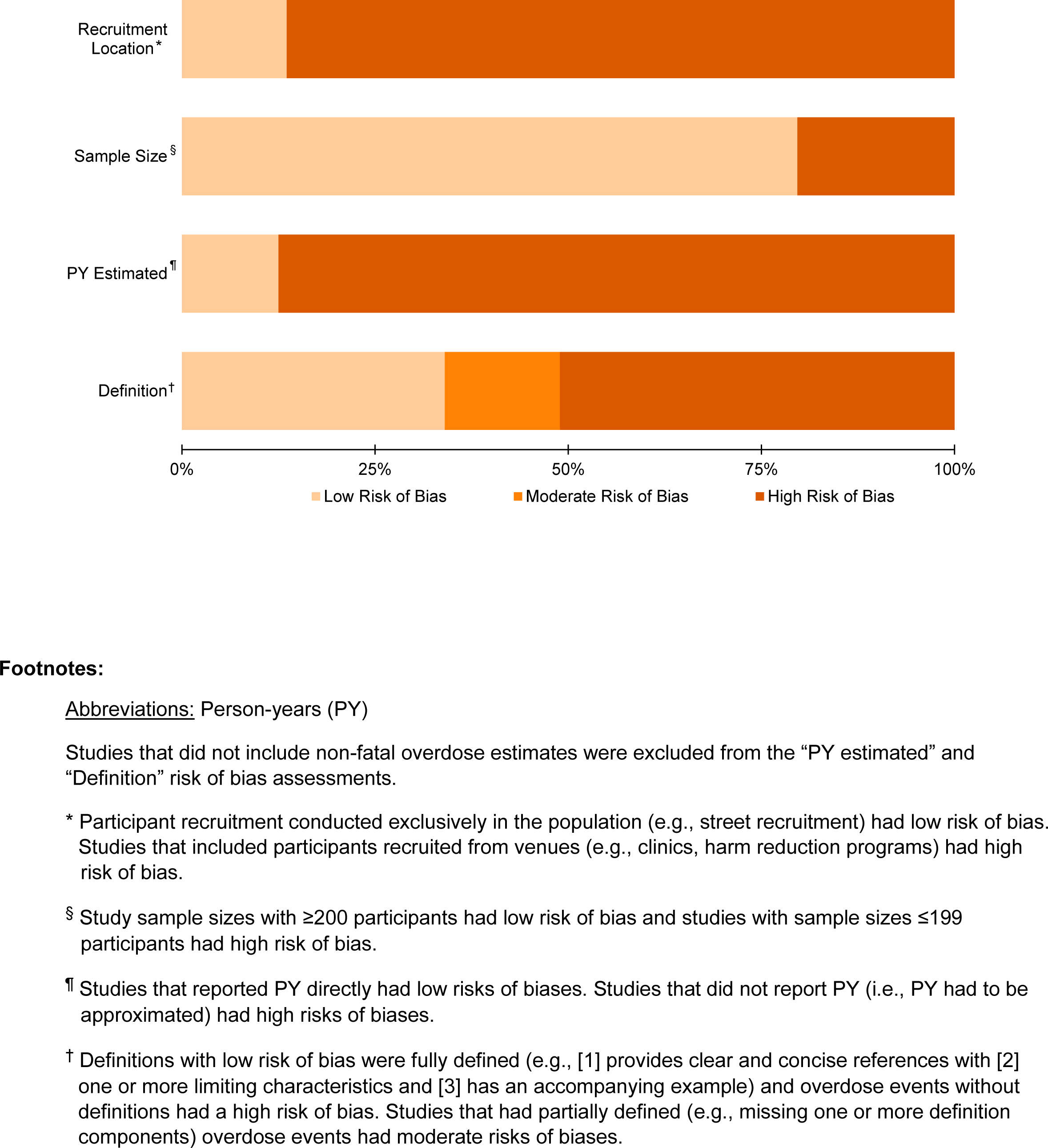
Risk of Bias Assessment for 58 Studies Included in the Meta-Analysis

### Statistical analysis

To ensure the comparability of metrics for non-fatal and fatal overdose, we calculated rates per person-year (PY) to estimate the occurrence of both types using the data from studies conducted at the same time period. Non-fatal and fatal overdose rates were calculated by dividing the number of events (i.e., non-fatal or fatal overdoses) by PY of follow-up in each study. For studies that did not report PY directly (e.g., cross-sectional studies), PY were approximated by multiplying the overdose recall period by the sample size (e.g., 100 people with a 6-month recall period contributes 50 PY). Reported or approximated PY were used as denominators for calculating non-fatal and fatal rates. We did not include non-fatal overdose estimates with fewer than three months of recall period, due to the likely flawed assumption that the frequency of events occurring in one, three-month period would be similar to all three-month periods during a given year. Study-level non-fatal and fatal rates are listed by country in Appendix 4.2 and depicted in forest plots sorted by median data collection year.

Despite 2013 often being described as the start of the third overdose wave, overdose deaths attributable to synthetic opioids increased dramatically after 2016 (CDC, 2023c; Mattson et al., 2021). From 2013 to 2017 synthetic opioid-involved overdose deaths increased 817%, with 2017 having the highest number of overdose deaths across all drug types in the previous decade (Casillas et al., 2024). To examine this later time period separately, we estimated non-fatal and fatal overdose rates using data collected exclusively after 2016. These estimates were meta-analyzed as a subset of, rather than a direct comparator to, estimates from the overall time period. This approach was chosen because many included studies collected data during time periods including years prior to and post-2016.

Overall rates of non-fatal and fatal overdose among PWID were pooled using random effects models to account for the variation in reporting methods, dates of data collection, and geographic location. The meta-analysis was conducted using the R program *meta (Balduzzi et al., 2019)*. Rates were log-transformed, and the random effects model was estimated using restricted maximum likelihood. Heterogeneity in the rates across studies were assessed using Tau^2^ and I^2^ (Michael Borenstein et al., 2009). Sensitivity analyses were conducted to explore probable sources of heterogeneity owing to differences in geographic location, dates of data collection, recruitment method, whether person-years were reported directly or estimated, and the survey recall period used to estimate person-years. Meta-regression was used to explore the extent to which heterogeneity was attributable to participant recruitment method, data collection year, or geographic location. Publication bias was examined using Egger’s test of funnel plot asymmetry (Egger et al., 1997). Ratios of non-fatal to fatal overdose rates were computed where possible given adequate data. To estimate confidence intervals for the ratios, we defined a distribution around each estimated rate using the mean and standard error. We resampled values from each rate distribution (i.e., non-fatal and fatal rates) and calculated resulting ratios (n=10,000). From the resulting distribution of ratios, we defined the 95% CI as the 2.5^th^ and 97.5^th^ percentiles.

## Results

### Search Results and Study Characteristics

The PRISMA flow diagram summarizes the study selection process (Figure 1). We screened 41,416 relevant titles and abstracts; 1,911 were selected for full report review. A total of 143 (7.5%) citations were included in the systematic review. The most common reasons for exclusion were having no overdose measure among PWID, reporting data exclusively collected before 2010, reporting overdose numbers without denominators, and reporting lifetime versus current measurements of IDU or non-fatal overdose.

**Figure 3.**
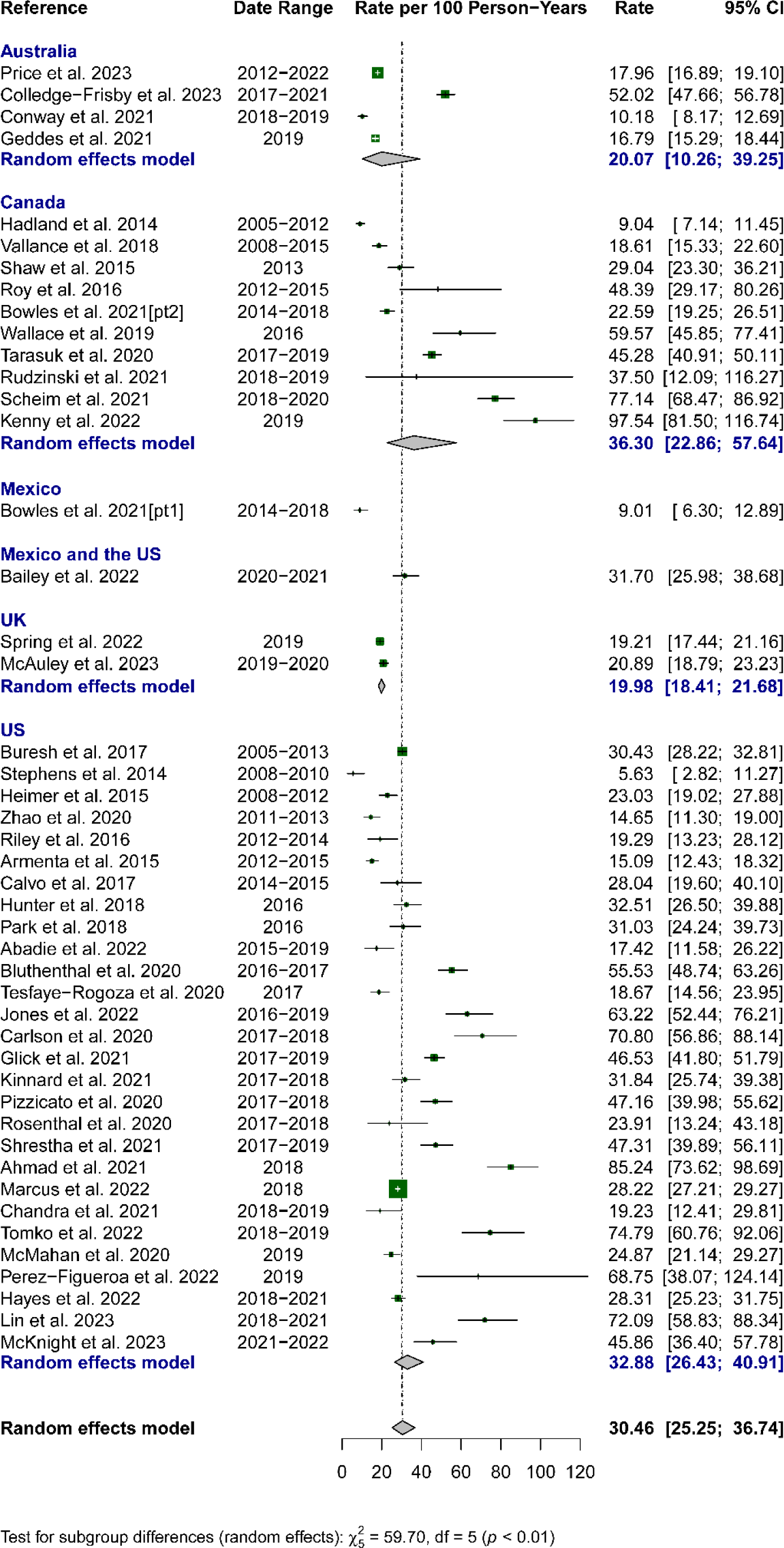
Random-Effects Meta-Analysis of Country-Level Non-Fatal Overdose Rates Per 100 PY

**Figure 4.**
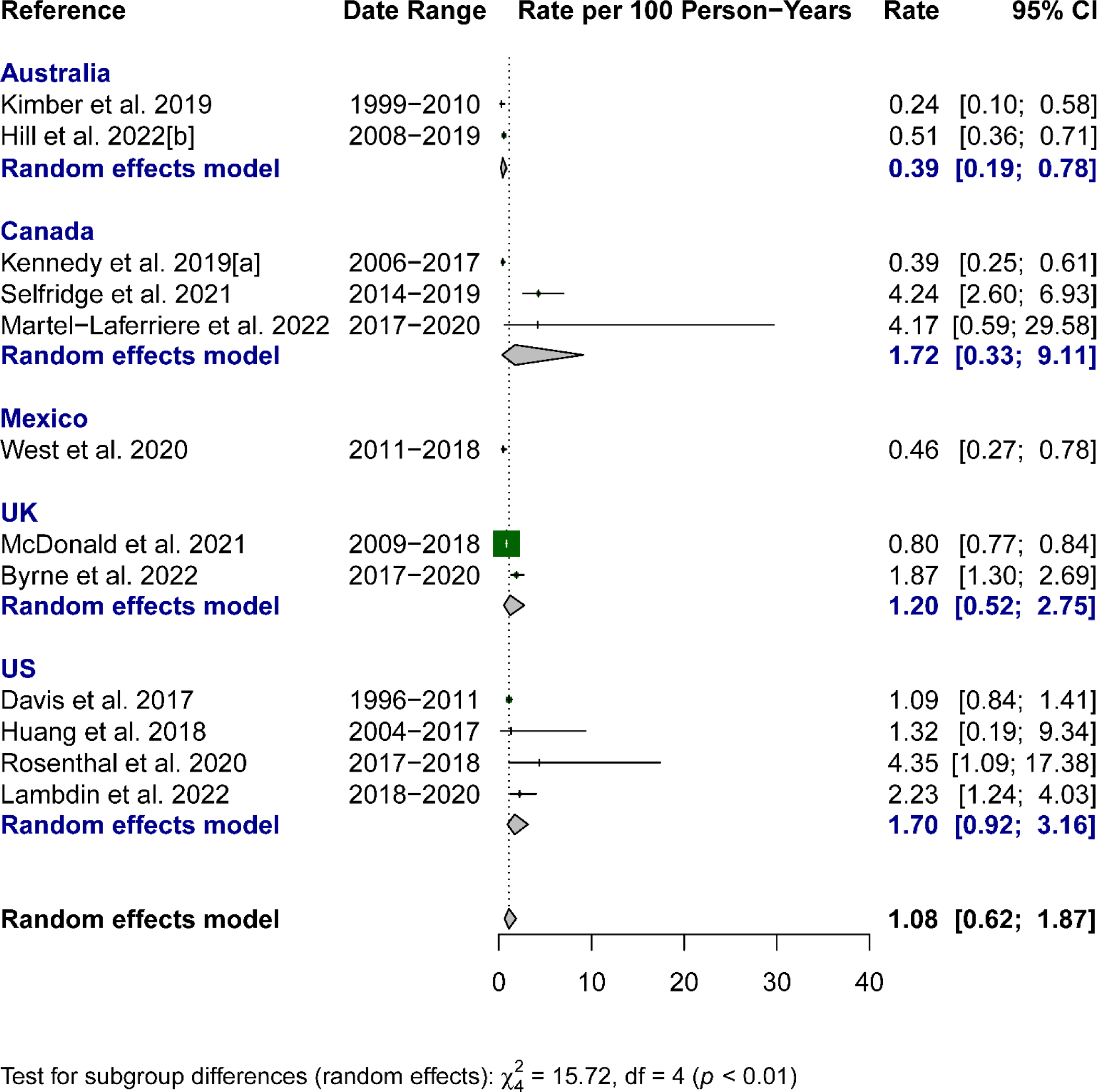
Random-Effects Meta-Analysis of Country-Level Fatal Overdose Rates Per 100 PY

Among 58 studies contributing to unique data to the meta-analysis, more than half of these studies were from the U.S. (53%), followed by Canada (24%), Australia (12%), the U.K. (7%), and Mexico (3%). Study sample sizes ranged from 15 – 35,065. The majority (86%) recruited participants using a combination of population and venue-based recruitment strategies (Figure 2) and 32% measured and reported overdose events longitudinally (Appendix 4.2). PY was estimated for 40 out of 46 studies (87%) that reported non-fatal overdose data. Among these 40 studies, 50% used 12-month recall periods, 45% used 6-month recall periods, and 5% used 3-month recall periods to approximate PY.

### Non-Fatal Overdose

Forty-six studies contributed data to the overall estimated non-fatal overdose rate, which showed 30.5 overdoses per 100 PY (95% CI: 25.3 – 36.7) (Table 1). Estimated non-fatal overdose rates were 32.5 per 100 PY (95% CI: 26.6 – 39.7; 40 studies) in North America, 32.9 per 100 PY (95% CI: 26.4 – 40.9; 28 studies) in the U.S., and 36.3 per 100 PY (95% CI: 22.9 – 57.6; 10 studies) in Canda. Comparatively, estimated non-fatal overdose rates were 20.0 per 100 PY (95% CI: 18.4 – 21.7; 2 studies) in the U.K. and 20.1 per 100 PY (95 CI: 10.3 – 39.3; 4 studies) in Australia.

**Table 1.** Non-Fatal and Fatal Overdose Rate Estimates (2010 – 2023)

| Location | N | Rate (95% CI)<sup>§</sup> | I<sup>2</sup> (%) | Tau<sup>2</sup> * | Ratio (NF:F) | Percent<sup>¶</sup> |
| --- | --- | --- | --- | --- | --- | --- |
| **Non-Fatal overdose** |  |  |  |  |  |  |
| Overall | 46 | 30.46 (25.25 – 36.74) | 98.00 | 0.40* |  |  |
| Post-2016<sup>†</sup> | 28 | 35.51 (28.05 – 44.96) | 98.30 | 0.38 |  |  |
| North America | 40 | 32.54 (26.64 – 39.74) | 97.20 | 0.39* |  |  |
| Post-2016<sup>†</sup> | 22 | 43.21 (35.05 – 53.28) | 97.50 | 0.23 |  |  |
| United States | 28 | 32.88 (26.43 – 40.91) | 96.60 | 0.33* |  |  |
| Post-2016<sup>†</sup> | 16 | 41.03 (32.07 – 52.50) | 97.20 | 0.23 |  |  |
| Canada | 10 | 36.30 (22.86 – 57.64) | 98.20 | 0.52* |  |  |
| Post-2016<sup>†</sup> | 5 | 55.32 (35.53 – 86.13) | 96.70 | 0.22 |  |  |
| Mexico | 1 | – | – | – |  |  |
| Post-2016<sup>†</sup> | – | – | – | – |  |  |
| Australia | 4 | 20.07 (10.26 – 39.25) | 99.40 | 0.46* |  |  |
| Post-2016<sup>†</sup> | 4 | 17.11 (7.88 – 37.12) | 99.30 | 0.62 |  |  |
| United Kingdom | 2 | 19.98 (18.41 – 21.68) | 23.60 | 0.00 |  |  |
| Post-2016<sup>†</sup> | 2 | 19.98 (18.41 – 21.68) | 23.60 | 0.00 |  |  |
| **Fatal Overdose** |  |  |  |  |  |  |
| Overall | 12 | 1.08 (0.62 – 1.87) | 90.70 | 0.77* | 28.20 (15.91 – 50.95) | 3.42% |
| Post-2016<sup>†</sup> | 5 | 1.80 (1.19 – 2.72) | 72.60 | 0.12 | 19.73 (12.39 – 31.83) | 4.82% |
| North America | 8 | 1.44 (0.70 – 2.97) | 90.20 | 0.85* | 22.60 (10.81 – 48.39) | 4.24% |
| Post-2016<sup>†</sup> | 3 | 2.57 (1.52 – 4.33) | 0.00 | 0.00 | 16.81 (9.69 – 29.71) | 5.61% |
| United States | 4 | 1.70 (0.92 – 3.16) | 62.20 | 0.21* | 19.34 (10.19 – 37.54) | 4.92% |
| Post-2016<sup>†</sup> | 2 | 2.47 (1.44 – 4.26) | 0.00 | 0.00 | 16.61 (9.24 – 30.28) | 5.68% |
| Canada | 3 | 1.72 (0.33 – 9.11) | 96.10 | 1.87* | 21.10 (3.88 – 121.31) | 4.52% |
| Post-2016<sup>†</sup> | 1 | – | – | – | – | – |
| Mexico | 1 | – | – | – | – | – |
| Post-2016<sup>†</sup> | – | – | – | – | – | – |
| Australia | 2 | 0.39 (0.19 – 0.78) | 59.00 | 0.16 | 51.46 (19.86 – 136.43) | 1.91% |
| Post-2016<sup>†</sup> | – | – | – | – | – | – |
| United Kingdom | 2 | 1.20 (0.52 – 2.75) | 95.10 | 0.34* | 16.65 (7.30 – 38.60) | 5.67% |
| Post-2016<sup>†</sup> | 2 | 1.44 (0.92 – 2.23) | 82.80 | 0.09 | 13.88 (8.92 – 21.88) | 6.72% |

Abbreviations: Sample Size (N); Confidence-Interval (CI); Person-years (PY); Non-Fatal (NF); Fatal (F)
Significance tests for the post-2016 analyses could not be calculated.
\* Q-test for heterogeneity p-value ≤ 0.05
§ Per 100 PY
¶ The percent of overdose events that result in death among PWID.
† Start dates for data collection began during or after 2017.

### Fatal Overdose

Among twelve studies contributing overdose mortality data, the overall estimated fatal overdose rate was 1.1 per 100 PY (95% CI: 0.6 – 1.9). The estimated fatal overdose rate in North America was 1.4 per 100 PY (95% CI: 0.7 – 3.0; 8 studies). There was little variation in estimated fatal overdose rates for the U.S. (1.7 fatal overdoses per 100 PY; 95% CI: 0.9 – 3.2; 4 studies) and Canada (1.7 per 100 PY; 95% CI: 0.3 – 9.1; 3 studies). Comparatively, Australia’s estimated fatal overdose rate was 0.4 per 100 PY (95% CI: 0.2 – 0.8; 2 studies) and the U.K.’s rate was 1.2 per 100 PY (95% CI: 0.5 – 2.8; 2 studies).

### Post-2016 Analysis

The overall estimated non-fatal overdose rate from data collected post-2016 was 35.5 (95% CI: 28.1 – 45.0; 28 studies), 17% higher than the 2010 – 2023 rate. The North American non-fatal overdose rate was 33% higher when sub-setting data to post-2016 (43.2 per 100 PY; 95% CI: 35.1 – 53.3; 22 studies). Non-fatal overdose rates in the U.S. were 25% and 52% higher, respectively, post-2016: 41.0 per 100 PY in the U.S. (95% CI: 32.1 – 52.5; 16 studies) and 55.3 per 100 PY in Canada (95% CI: 35.5 – 86.1; 5 studies). Non-fatal overdose rates in Australia post-2016 (17.1 per 100 PY; 95% CI: 7.9 – 37.1; 4 studies) were 15% lower than over the full time period. There were no changes in the U.K.’s post-2016 rate from the 2010 – 2023 rate.

In North America, the rates for fatal overdose were even more pronounced, when sub- setting data to post-2016 rate. The fatal overdose rate was 80% higher post-2016 (2.6 per 100 PY; 95% CI: 1.5 – 4.3; 3 studies) compared to 1996 – 2022. Country-specific post-2016 estimates for non-fatal overdose could only be made for the U.S. (2.5 per 100 PY; 95% CI: 1.4 – 4.3) and the U.K. (1.4 per 100 PY; 95% CI: 0.9 – 2.2), but each are based on only two studies.

### Ratios of non-fatal to fatal overdose rates

Overall, the estimated ratio of non-fatal to fatal overdose events for pooled estimates was 28.2 (CI: 15.9 – 51.0):1, meaning 3.4% of estimated overdoses among PWID were fatal, compared to 19.7 (CI: 12.4 – 31.8):1 (4.8% of overdoses were fatal) when limiting estimates to those derived exclusively from post-2016 estimates. In the U.S., the estimated ratio of non-fatal to fatal overdose events for pooled estimates was 19.3 (CI: 10.2 – 37.5):1, indicating approximately 5% of overdoses among PWID were fatal. In contrast, when focusing solely on post-2016 estimates, the ratio was 16.6 (CI: 9.2 – 30.3):1, with 5.7% of overdoses being fatal. The ratio was considerably higher in Australia (51.5 (CI: 19.9 – 136.4):1) than in other countries, where ratios were similar to one another, overall and post-2016.

### Heterogeneity and Risk of Bias Assessments

Substantial heterogeneity was observed across both non-fatal and fatal overdose estimates, including country-specific estimates (Table 1). To determine the extent to which observed heterogeneity in non-fatal overdose estimates were associated with research design differences, we compared estimates by recruitment and overdose measurement methods (Appendix 4.1). Non-fatal overdose rates were significantly higher among studies for which we approximated person-year denominators compared to those reporting overdose rates directly (32.9 versus 17.8, p<0.01). In a meta-regression model, studies with six- or three-month recall periods had significantly higher non-fatal overdose rates compared to those with a 12-month recall period. In the U.S., no significant differences in non-fatal overdose rates were found among PWID recruited from the population (e.g., street recruitment, respondent-driven sampling, etc.), venues (e.g., harm reduction programs, clinics, etc.), or using a combination of population- and venue-based recruitment. In a meta-regression model controlling for recruitment location, time (data exclusively collected post-2016 versus earlier) significantly contributed to observed heterogeneity in non-fatal overdose estimates (data not shown in table). There were an insufficient number of studies providing fatal overdose estimates to explore sources of heterogeneity. To understand the effect extreme study values had on pooled rate estimates, sensitivity analyses removing studies with post-2016 non-fatal overdose rates 120% above the mean estimate were conducted. Removing Kenny et al. (2022) reduced Canada’s post-2016 non-fatal overdose rate by 17% and removing Ahmad et al. (2021) reduced the U.S. rate by 5% (data not shown).

Egger’s test of funnel plot asymmetry revealed no evidence of publication bias for the overall or post-2016 non-fatal overdose rates (data not shown). There were insufficient numbers of studies reporting fatal overdose rates post-2016 to examine funnel plot asymmetry.

## Discussion

Findings from this systematic review and meta-analysis demonstrate substantial burden of non-fatal and fatal overdose among PWID in the U.S. and comparator countries. Post-2016 pooled estimates of overdose among PWID are the first containing data exclusive to the third wave of the opioid epidemic. Additionally, these findings provide a novel understanding of the relationship between non-fatal and fatal overdose among PWID. Using the post-2016 pooled rates presented here, we estimate there were approximately 17 non-fatal overdoses for every fatal overdose experienced by PWID living in the U.S., meaning an estimated 1/18 (6%) of all overdoses experienced by PWID in the U.S. resulted in death after 2016.

The ratio of non-fatal to fatal overdose provides critical information on the burden of largely preventable death conditional on overdose (Casillas et al., 2024). When monitored over time and across geographies, this metric can measure success of public health interventions in terms of mortality prevention. In the U.S., the overall overdose mortality rate quadrupled from 2010 – 2022 (Spencer et al., 2024), while non-fatal overdose events increased by 19% from 2010 – 2020 (Casillas et al., 2024). The larger increase in mortality relative to overdose events means that, among PWUD, overdoses are becoming increasingly fatal. Casillas et al. recently demonstrated the ratio of non-fatal to fatal overdose among PWUD halved in the U.S. from 20.4 in 2010 to 10.1 in 2020. Increased lethality of overdose is due in large part to changes in the U.S. drug supply toward more lethal substances (e.g., xylazine, synthetic opioids) (California Department of Public, 2024; CDC, 2023b, 2024f; DEA, 2022) and increased polysubstance use including combinations of synthetic opioids and stimulants (O’Donnell et al., 2021).

Regulating the drug supply is largely outside the purview of public health, apart from provision of drug testing supplies and services; however, prevention of mortality among persons experiencing overdose is squarely in the purview of public health. Mortality prevention should be prioritized by public health practitioners given increases in mortality relative to overdose among PWUD and the consistently high rates of mortality relative to overdose shown here among PWID. Evidence-based harm reduction interventions such as naloxone provision and potentially SCSs are effective in preventing overdose mortality by counteracting the effects of opioids and their analogs and shortening response times to overdose events (Kennedy, Hayashi, et al., 2019; McDonald & Strang, 2016). Substance use disorder treatment, including MOUD, is also effective in preventing non-fatal and fatal overdoses by reducing exposure to lethal substances (Strange et al., 2022) but is only an option for persons with treatment readiness. Harm reduction interventions to prevent non-opioid overdose mortality for PWUD including PWID are critically needed.

While we provide time-bound, geographically-specific estimates of the ratio of non-fatal to fatal overdose among PWID, it will be important to monitor the relationship between non-fatal and fatal overdose over time and across geographies. While Casillas and colleagues estimated this ratio over time for PWUD using routinely collected surveillance data, they were unable to classify overdose events by route of administration (Casillas et al., 2024). This is a current problematic limitation of overdose surveillance data, which do not routinely collect information on mode of substance use due largely to substantial logistical difficulties of doing so (CDC, 2023a, 2024a, 2024d). Currently, NHBS is the only source of U.S. surveillance data on non-fatal overdose among PWID (CDC, 2024c, 2024e). However, due to its tri-annual cycle, underrepresentation of PWID living in rural communities, measurement of opioid overdoses only, and lack of fatal overdose measurement, NHBS provides only partial information needed for a comprehensive understanding of overdose among PWID (CDC, 2024c, 2024e; Lerner & Fauci, 2019). NHBS data indicate 28% and 27% of PWID experienced at least one non-fatal opioid overdose in 2018 and 2022, respectively. It is unclear how these estimates compare to the 41/100PY estimated here, because they represent only opioid overdoses in select cities and do not measure the number of overdoses experienced by a single person during a 12-month period.

Due to these surveillance limitations, meta-analysis of research study data is an important tool for estimating the burden of overdose, and the relationship between non-fatal and fatal overdose, among PWID. To robustly produce these estimates over time, on-going and current estimates of overdose are needed from research studies, particularly for fatal overdose, for which many fewer estimates are currently available relative to non-fatal overdose. To facilitate future meta-analyses, we recommend standardized collection of overdose indicators across PWID-focused research studies. First, to accurately assess a current rate of non-fatal overdose, research studies should consider measuring the number of overdose and, separately, overamping (e.g., an overdose resulting from the use of stimulant drugs such as speed, cocaine, or meth) events over a defined recall period. Many studies we excluded reported overdose among people who had “ever” injected drugs or measured “ever” overdosing among people who recently injected drugs. Neither of these measurements provide an indicator of current overdose risk among PWID. For studies that measured “ever” overdosing during a defined time period, we assumed equal risk for overdose over such time periods up to 12 months in our approximation of person-time. To compute non-fatal overdose rates that can be directly compared to fatal overdose rates, the actual number of non-fatal overdoses experienced is needed. Second, we recommend that studies including both former and recent PWID stratify overdose estimates by recency of injection behavior and by demographic characteristics where possible. Data sparsity inhibited us from producing stratified estimates by gender, age, and race. Recent research on PWUD demonstrates differential overdose risk by gender, age, and race, making these stratifications critical (Britz et al., 2023; Spencer et al., 2022; Zang et al., 2023). Third, we recommend studies report overall and cause-specific mortality rates from death registry matches where possible, potentially as part of loss to follow up analyses. If uniformly applied, these recommendations could substantially increase the quality of overdose data among PWID and information available for meta-analyses. Additionally, increased collaboration and data sharing among researchers and service providers focused on PWID health has potential to increase timeliness of pooled estimates (Bradley et al., 2022).

Generally, more PWID-focused research is needed to accurately measure overdose rates in under-represented populations, particularly in rural areas. The national estimates presented here are a step forward in understanding current overdose risk among PWID, but more geographically granular estimates are needed to guide local resource allocation and interventional strategies. Some states or cities may have local information needed to produce the ratio of non-fatal to fatal overdose among PWID, which includes the number of non-fatal and fatal overdoses experienced by PWID, and either person years of observation or PWID population size during a given time period. Some of these indicators may be drawn from local research studies or surveillance data, but the lack of local PWID population size estimates remain a major limitation in most U.S. geographic areas.

In addition to developing a metric for understanding the burden of mortality conditional on overdose among PWID, these findings provide updates to previous estimates of non-fatal overdose among PWID in the U.S. and four other countries. Degenhardt and colleagues (2023) recently published global estimates of non-fatal overdose prevalence among PWID by country. Pooled estimates from this previous review included data through April 2022 but also included estimates from their previous reviews dating back to 2007, with no exclusions distinguishing between when data were collected for reports versus published in reports Because these estimates are derived from more recent data collections, and rates rather than percentages were reported, they are substantially higher than estimates in the Degenhardt et al. review. For example, the post-2016 U.S. non-fatal overdose rate in our review was 41.0/100 person-years compared to 19.4% in the previous review (Degenhardt et al., 2023). The post-2016 non-fatal overdose rate in Australia was 17.1/100 person-years compared to 8.5% in the previous review. There are also some similarities between findings from these reviews, including the finding that PWID living in Canada had the highest burden of non-fatal overdose in North America. These findings also differ from a review of data collected between 2006 – 2016, which, for example, reported similar non-fatal overdose rates for Canada and the U.S. (Colledge et al., 2019). To our knowledge, no other systematic review or meta-analysis has estimated pooled rates for overdose mortality among PWID.

Changes in overdose rates over time are likely attributable to a combination of shifts in the drug supply toward more lethal substances and change in availability of harm reduction services, including possible disruption of services during the COVID-19 pandemic (Bailey et al., 2023; Kariisa et al., 2023; Tanz et al., 2023). In this review, PWID in Canada and the U.S. had the highest rates of both non-fatal and fatal overdose, potentially reflecting overlaps in drug supply. The UK and Australia had substantially lower rates of non-fatal overdose, and Australia had the lowest overdose mortality rate. Contrary to what was expected, the post-2016 non-fatal overdose rate in Australia was lower than the overall rate, which may be due to widespread implementation of overdose prevention and harm reduction interventions including SCSs (Colledge-Frisby et al., 2023; Sordo et al., 2017; Victoria, 2020). Canada, the U.K., and Australia provide naloxone over the counter and widely distribute kits in communities, a practice recently adopted by the U.S. (FDA, 2023; Seo et al., 2024; WHO, 2023). Additional resources are urgently needed for scale up of evidence-based interventions (e.g., community-based naloxone distribution, lower barrier access to MOUD) that prevent overdose mortality in the U.S. given recent increases in the PWID population (Bradley et al., 2023) and large increases in overdose mortality risk over time (Casillas et al., 2024; CDC, 2024b).

## Limitations

In sensitivity analyses, we identified multiple sources of heterogeneity in the pooled overdose estimates. These included calendar time, whether person-time was reported directly by the study or estimated by the study team, and length of the recall period for studies not reporting person-time. Heterogeneity was reduced for post-2016 estimates compared to those spanning the total time period, but data sparsity limited geographically-specific, post-2016 estimates for some countries. Differences in estimates from studies reporting versus not reporting person-time may be attributable to two factors. First, computing person time from the study recall period requires the assumption that overdose risk in one 3- or 6-month period was transferrable to other 3- or 6-month periods across 12-months. If persons experiencing overdose during a <12 month recall period did not experience repeat overdoses across 12 months, the overdose rate will be over-estimated. On the other hand, if persons reporting “any overdose” in a 12-month period experienced multiple overdoses during this time, the overdose rate will be under-estimated. Second, many studies directly reporting PY were from the U.S. with data collected prior to 2016. Data sparsity limited our ability to simultaneously control for calendar time and reporting of person-time. Among U.S. studies, we did not find significant differences in overdose rates among PWID recruited from the population versus venues, likely due to limited power to do so. There were not enough studies to make this assessment for other countries. Despite these sensitivity analyses, substantial heterogeneity remains in many of the estimates reported here, likely due to within-country geographic variability and differences in measurement and reporting addressed previously.

This review is subject to several limitations. First, we restricted our review to studies published in English. In consideration of this limitation, we cross-referenced our references with that of a global systematic review on PWID that extensively searched unpublished literature for overdose estimates (Degenhardt et al., 2023). There was only one unpublished report with data on recent overdose among PWID living in the U.S. that was not included in our review. Second, as discussed previously, available data on non-fatal and fatal overdose among PWID varied by recruitment site, geographic location, time period, and risk population (e.g., persons with HIV, HCV) resulting in significant heterogeneity in pooled overdose estimates. Third, pooled overdose rate estimates are sensitive to extreme values, particularly from studies with small sample sizes. Excluding these extreme values in a post-2016 sensitivity analyses resulted in a decline in non-fatal overdose rates for Canada and the U.S. Finally, to produce effect sizes with comparable metrics (i.e., rates per 100 PY) we expressed non-fatal overdose as a rate as opposed to a proportion. This may have under- or over-estimated overdose rates as previously discussed.

## Conclusion

PWID have extremely high risk for overdose, and we estimate 5% of overdoses among PWID in the U.S. resulted in death after 2016. This percentage can be considerably reduced through investments in evidence-based harm reduction programs, and by prioritizing interventions that prevent mortality (e.g., naloxone, MOUD, and promotion of safer injection practices). Because current U.S. surveillance systems are unable to provide data on route of administration, research studies remain the most important sources of data on PWID health outcomes. Standardizing non-fatal overdose reporting measurement and reporting methods would facilitate greatly improved quality and precision of overdose burden estimates. With improvements in overdose reporting methods, future meta-analyzed overdose estimates can evaluate the success of intervention strategies over time and promote resource allocation based on current needs.

## Data Availability

All data produced in the present study are available upon reasonable request to the authors

## Acknowledgements

The authors acknowledge Shenita Peterson for assistance with our search strategy and Marcus Goff and Jonathan Standish for assistance with data extraction. The authors acknowledge funding for this work from the Centers for Disease Control and Prevention, National Center for National Center for HIV, Viral Hepatitis, STD, and TB Prevention (U38PS004650) and the National Institutes of Health, National Institute on Drug Abuse (R01DA051302).

