## Supplemental materials for "Systematic review and meta-analysis to estimate the burden of non-fatal and fatal overdose among people who inject drugs living in the U.S. and comparator countries: 2010 – 2023"

### **Appendix 1:** PRISMA Checklist

| Section and Topic | Item # | Checklist item | Location where item is reported |
|---|---|---|---|
| **TITLE** | | | |
| Title | 1 | Identify the report as a systematic review. | Pg. 1 |
| **ABSTRACT** | | | |
| Abstract | 2 | See the PRISMA 2020 for Abstracts checklist. | Pg. 2 |
| **INTRODUCTION** | | | |
| Rationale | 3 | Describe the rationale for the review in the context of existing knowledge. | Pg. 4 |
| Objectives | 4 | Provide an explicit statement of the objective(s) or question(s) the review addresses. | Pg. 4 |
| **METHODS** | | | |
| Eligibility criteria | 5 | Specify the inclusion and exclusion criteria for the review and how studies were grouped for the syntheses. | Pg. 5; Fig. 1 |
| Information sources | 6 | Specify all databases, registers, websites, organisations, reference lists and other sources searched or consulted to identify studies. Specify the date when each source was last searched or consulted. | Pg. 5; App. 2 |
| Search strategy | 7 | Present the full search strategies for all databases, registers and websites, including any filters and limits used. | App. 2 |
| Selection process | 8 | Specify the methods used to decide whether a study met the inclusion criteria of the review, including how many reviewers screened each record and each report retrieved, whether they worked independently, and if applicable, details of automation tools used in the process. | Pg. 5-6 |
| Data collection process | 9 | Specify the methods used to collect data from reports, including how many reviewers collected data from each report, whether they worked independently, any processes for obtaining or confirming data from study investigators, and if applicable, details of automation tools used in the process. | Pg. 6 |
| Data items | 10a | List and define all outcomes for which data were sought. Specify whether all results that were compatible with each outcome domain in each study were sought (e.g. for all measures, time points, analyses), and if not, the methods used to decide which results to collect. | Pg. 6 |
| | 10b | List and define all other variables for which data were sought (e.g. participant and intervention characteristics, funding sources). Describe any assumptions made about any missing or unclear information. | Pg. 6 |
| Study risk of bias assessment | 11 | Specify the methods used to assess risk of bias in the included studies, including details of the tool(s) used, how many reviewers assessed each study and whether they worked independently, and if applicable, details of automation tools used in the process. | Pg. 6-7; Fig. 2; App. 3 |
| Effect measures | 12 | Specify for each outcome the effect measure(s) (e.g. risk ratio, mean difference) used in the synthesis or presentation of results. | Pg. 6-7 |
| Synthesis methods | 13a | Describe the processes used to decide which studies were eligible for each synthesis (e.g. tabulating the study intervention characteristics and comparing against the planned groups for each synthesis (item #5)). | Pg. 6-7 |
| | 13b | Describe any methods required to prepare the data for presentation or synthesis, such as handling of missing summary statistics, or data conversions. | Pg. 6-7 |
| | 13c | Describe any methods used to tabulate or visually display results of individual studies and syntheses. | Pg. 6-7 |
| | 13d | Describe any methods used to synthesize results and provide a rationale for the choice(s). If meta-analysis was performed, describe the model(s), method(s) to identify the presence and extent of statistical heterogeneity, and software package(s) used. | Pg. 6-7 |
| | 13e | Describe any methods used to explore possible causes of heterogeneity among study results (e.g. subgroup analysis, meta-regression). | Pg. 6-7 |
| | 13f | Describe any sensitivity analyses conducted to assess robustness of the synthesized results. | Pg. 6-7 |
| Reporting bias assessment | 14 | Describe any methods used to assess risk of bias due to missing results in a synthesis (arising from reporting biases). | Pg. 6-7 |
| Certainty assessment | 15 | Describe any methods used to assess certainty (or confidence) in the body of evidence for an outcome. | N/a |

| Section and Topic | Item # | Checklist item | Location where item is reported |
|---|---|---|---|
| **RESULTS** | | | |
| Study selection | 16a | Describe the results of the search and selection process, from the number of records identified in the search to the number of studies included in the review, ideally using a flow diagram. | Pg. 8 |
| | 16b | Cite studies that might appear to meet the inclusion criteria, but which were excluded, and explain why they were excluded. | N/a |
| Study characteristics | 17 | Cite each included study and present its characteristics. | App. 4.2 |
| Risk of bias in studies | 18 | Present assessments of risk of bias for each included study. | App.3 |
| Results of individual studies | 19 | For all outcomes, present, for each study: (a) summary statistics for each group (where appropriate) and (b) an effect estimate and its precision (e.g. confidence/credible interval), ideally using structured tables or plots. | Fig. 3-4; App. 4.2; 6.1-6.2 |
| Results of syntheses | 20a | For each synthesis, briefly summarise the characteristics and risk of bias among contributing studies. | Fig. 2; App. 3 |
| | 20b | Present results of all statistical syntheses conducted. If meta-analysis was done, present for each the summary estimate and its precision (e.g. confidence/credible interval) and measures of statistical heterogeneity. If comparing groups, describe the direction of the effect. | Pg. 8-10; Tbl. 1; App. 4.1 |
| | 20c | Present results of all investigations of possible causes of heterogeneity among study results. | Pg.9-10; App. 4.1 |
| | 20d | Present results of all sensitivity analyses conducted to assess the robustness of the synthesized results. | Pg. 9-10; App. 4.1 |
| Reporting biases | 21 | Present assessments of risk of bias due to missing results (arising from reporting biases) for each synthesis assessed. | Pg. 9-10 |
| Certainty of evidence | 22 | Present assessments of certainty (or confidence) in the body of evidence for each outcome assessed. | N/a |
| **DISCUSSION** | | | |
| Discussion | 23a | Provide a general interpretation of the results in the context of other evidence. | Pg. 10-14 |
| | 23b | Discuss any limitations of the evidence included in the review. | Pg. 14-15 |
| | 23c | Discuss any limitations of the review processes used. | Pg. 14-15 |
| | 23d | Discuss implications of the results for practice, policy, and future research. | Pg. 10-14 |
| **OTHER INFORMATION** | | | |
| Registration and protocol | 24a | Provide registration information for the review, including register name and registration number, or state that the review was not registered. | N/a |
| | 24b | Indicate where the review protocol can be accessed, or state that a protocol was not prepared. | N/a |
| | 24c | Describe and explain any amendments to information provided at registration or in the protocol. | N/a |
| Support | 25 | Describe sources of financial or non-financial support for the review, and the role of the funders or sponsors in the review. | Pg. 2, 15 |
| Competing interests | 26 | Declare any competing interests of review authors. | N/a |
| Availability of data, code and other materials | 27 | Report which of the following are publicly available and where they can be found: template data collection forms; data extracted from included studies; data used for all analyses; analytic code; any other materials used in the review. | N/a |

*From:* Page MJ, McKenzie JE, Bossuyt PM, Boutron I, Hoffmann TC, Mulrow CD, et al. The PRISMA 2020 statement: an updated guideline for reporting systematic reviews. BMJ 2021;372:n71. doi: 10.1136/bmj.n71

For more information, visit: http://www.prisma-statement.org/

### **Appendix 2:** Search Strategy

| *PubMed* |
|---|
| overdose PWID |
| overdose IDU |
| fatal* PWID |
| fatal* IDU |
| mortal* PWID |
| mortal* IDU |
| overdose inject* AND drug |
| (((overdose or fatal* or mortal*) AND (inject* or PWID or IDU)) AND (drug* or narcotic)) AND (NOT rat or mouse or mice)) |
| ((((((((((overdose or fatal or mortal) AND (inject or PWID or IDU)) AND (drug or narcotic))) ) AND (("2010"[Date - Publication] : "3000"[Date - Publication]))) NOT (rat)) NOT (mouse)) NOT (mice)) NOT (dog)) NOT (horse) |
| ("people who inject drugs"[tw] OR PWID*[tw] OR "IV Drug User*" OR "Intravenous Drug User*" OR (Inject* drug use*[tw]) OR IDU[tw] OR "Substance Abuse, Intravenous"[Mesh] OR "Needle Sharing"[Mesh]) AND (Overdose*[tw]) AND (Fatal*[tw] OR Mortal*[tw] OR Death[tw] OR Surviv*[tw]) AND English[lang] AND ("2010"[Date - Publication] : "2023"[Date - Publication]) NOT ("animals"[MeSH Terms] NOT "humans"[MeSH Terms]) |
| *Embase* |
| overdose pwid' OR (('overdose'/exp OR overdose) AND pwid) |
| overdose AND idu |
| fatal* AND PWID |
| fatal* AND idu |
| mortal* AND pwid |
| mortal* IDU |
| overdose AND inject* AND drug |
| ((('drug overdose'/exp OR 'drug overdose' OR fatal* OR mortal*:af) AND inject* OR pwid OR 'idu'/exp OR idu) AND drug* OR narcotic) NOT ('rat'/exp OR rat) NOT ('mouse'/exp OR mouse) NOT ('mice'/exp OR mice) NOT ('dog'/exp OR dog) NOT ('horse'/exp OR horse) NOT ('animal'/exp OR animal) AND [1-1-2010]/sd NOT [29-9-2023]/sd |
| "(""people who inject drugs"":ti,ab,kw,de OR PWID*:ti,ab,kw,de OR ""IV Drug User*"":ti,ab,kw,de OR ""Intravenous Drug User*"":ti,ab,kw,de OR (Inject* drug use*:ti,ab,kw,de) OR IDU:ti,ab,kw,de OR 'substance abuse'/exp OR 'needle sharing'/exp) AND (Overdose*:ti,ab,kw,de) AND (Fatal*:ti,ab,kw,de OR Mortal*:ti,ab,kw,de OR Death:ti,ab,kw,de OR Surviv*:ti,ab,kw,de) AND [english]/lim AND [2010-2023]/py NOT [animals]/lim" |
| *PsycINFO* |
| overdose PWID |
| overdose IDU |
| fatal* PWID |
| fatal* IDU |
| mortal* PWID |
| mortal* IDU |
| overdose inject* AND drug |
| ( overdose or poisoning or toxicity ) AND ( fatal or injury ) AND ( mortality or mortality rate or death or death rate ) AND inject* OR ( pwid or idu or injection drug users or injection drug addicts or people who inject drugs ) AND drug use OR narcotics NOT ( case report or case study or clinical case ) NOT rats NOT ( mouse or mice ) |
| (TX ( "people who inject drugs" OR PWID* OR "IV Drug User*" OR "Intravenous Drug User*" OR IDU ) OR TX (Inject* drug use*) OR (DE "Intravenous Drug Usage" OR DE "Needle Sharing")) AND TX (Overdose*) AND TX (Fatal* OR Mortal* OR Death OR Surviv*) AND (ZL "english") AND (DT 20100101-20230929) NOT (ZP "animal") |
| *ProQuest* |
| (overdose pwid) AND (la.exact("ENG") AND pd(20100101-20230929)) |
| (overdose idu) AND (la.exact("ENG") AND pd(20100101-20230929)) |
| (fatal pwid) AND (la.exact("ENG") AND pd(20100101-20230929)) |
| (fatal idu) AND (la.exact("ENG") AND pd(20100101-20230929)) |
| (mortal pwid) AND (la.exact("ENG") AND pd(20100101-20230929)) |
| (mortal idu) AND (la.exact("ENG") AND pd(20100101-20230929)) |
| (Overdose inject AND drug) AND (la.exact("ENG") AND pd(20100101-20230929)) |
| (((overdose OR poisoning OR toxicity) AND (fatal OR injury) AND (mortality OR mortality rate OR death OR death rate) AND inject* OR (pwid OR idu OR injection drug users OR injection drug addicts OR people who inject drugs) AND drug use OR narcotics NOT (case report OR case study OR clinical case) NOT rats NOT (mouse OR mice OR animal OR rat OR dog OR fish)) |
| noft(("people who inject drugs" OR PWID* OR "IV Drug User*" OR "Intravenous Drug User*" OR (Inject* drug use*) OR IDU) AND (Overdose*) AND (Fatal* OR Mortal* OR Death OR Surviv*)) |

Publication Year Limit: January 1, 2010 to September 29, 2023 (last searched September 29, 2023)

### **Appendix 3:** Risk of Bias Assessment Scores for Studies Included in the Meta-Analysis (N=58)

| Reference | Location | Recruitment Site | Sample Size | PY Estimated | Definition |
|---|---|---|---|---|---|
| College-Frisbey et al. 2023 | Australia | ⊗ | ⊕ | ⊗ | ⊗ |
| Conway et al., 2021 | Australia | ⊗ | ⊕ | ⊗ | ⊗ |
| Geddes et al., 2021 | Australia | ⊗ | ⊕ | ⊗ | ⊗ |
| Hill et al., 2022b | Australia | ⊗ | ⊕ | ○ | ○ |
| Kimber et al., 2019 | Australia | ⊗ | ⊕ | ○ | ○ |
| Price et al., 2023 | Australia | ⊗ | ⊕ | ⊗ | ⊕ |
| VanDenBoom et al., 2021 | Australia | ⊗ | ⊕ | ⊗ | ⊗ |
| Bowles et al., 2021[₪] | Canada | ⊗ | ⊕ | ⊗ | ⊕ |
| Hadland et al., 2014 | Canada | ⊗ | ⊕ | ⊕ | ⊕ |
| Kennedy et al., 2019a | Canada | ⊗ | ⊕ | ○ | ○ |
| Kenny et al., 2022 | Canada | ⊗ | ⊕ | ⊗ | ⊕ |
| Martel-Laferriere et al., 2022 | Canada | ⊗ | ⊗ | ○ | ○ |
| McCrae et al., 2020 | Canada | ⊗ | ⊕ | ⊗ | ⊕ |
| Roy et al., 2016 | Canada | ⊗ | ⊗ | ⊗ | ⊗ |
| Rudzinski et al., 2021 | Canada | ⊗ | ⊗ | ⊗ | ⊗ |
| Scheim et al., 2021 | Canada | ⊗ | ⊕ | ⊗ | ⊖ |
| Selfridge et al., 2021 | Canada | ⊗ | ⊕ | ○ | ○ |
| Shaw et al., 2015 | Canada | ⊕ | ⊕ | ⊗ | ⊗ |
| Tarasuk et al., 2020 | Canada | ⊗ | ⊕ | ⊗ | ⊗ |
| Vallance et al., 2018 | Canada | ⊗ | ⊕ | ⊗ | ⊗ |
| Wallace et al., 2019 | Canada | ⊗ | ⊗ | ⊗ | ⊕ |
| Bowles et al., 2021[₪] | Mexico | ⊗ | ⊕ | ⊗ | ⊕ |
| West et al., 2020 | Mexico | ⊗ | ⊕ | ○ | ○ |
| Bailey et al., 2022 | NA | ⊗ | ⊕ | ⊗ | ⊕ |
| Byrne et al., 2022 | UK | ⊗ | ⊕ | ○ | ○ |
| McAuley et al., 2023 | UK | ⊗ | ⊕ | ⊗ | ⊖ |
| McDonald et al., 2021 | UK | ⊗ | ⊕ | ○ | ○ |
| Spring et al., 2022 | UK | ⊗ | ⊕ | ⊗ | ⊖ |
| Abadie et al., 2022 | US | ⊗ | ⊗ | ⊕ | ⊗ |
| Ahmad et al., 2021 | US | ⊗ | ⊕ | ⊗ | ⊖ |
| Armenta et al., 2015 | US | ⊗ | ⊕ | ⊕ | ⊕ |
| Bluthenthal et al., 2020 | US | ⊗ | ⊕ | ⊗ | ⊗ |
| Buresh et al., 2017 | US | ⊕ | ⊕ | ⊕ | ⊕ |
| Calvo et al., 2017 | US | ⊗ | ⊗ | ⊗ | ⊖ |
| Carlson et al., 2020 | US | ⊕ | ⊕ | ⊗ | ⊗ |
| Chandra et al., 2021 | US | ⊗ | ⊗ | ⊗ | ⊗ |
| Davis et al., 2017 | US | ⊕ | ⊕ | ○ | ○ |
| Glick et al., 2021 | US | ⊗ | ⊕ | ⊗ | ⊕ |

| Reference | Location | Recruitment Site | Sample Size | PY Estimated | Definition |
|---|---|---|---|---|---|
| Hayes et al., 2022 | US | ⊗ | ⊕ | ⊗ | ⊗ |
| Heimer et al., 2015 | US | ⊗ | ⊕ | ⊗ | ⊗ |
| Huang et al., 2018 | US | ⊗ | ⊗ | ○ | ○ |
| Hunter et al., 2018 | US | ⊗ | ⊕ | ⊗ | ⊗ |
| Jones et al., 2022 | US | ⊕ | ⊕ | ⊗ | ⊗ |
| Kinnard et al., 2021 | US | ⊗ | ⊕ | ⊗ | ⊗ |
| Lambdin et al., 2022 | US | ⊗ | ⊗ | ○ | ○ |
| Lin et al., 2023 | US | ⊗ | ⊕ | ⊗ | ⊗ |
| Marcus et al., 2022 | US | ⊕ | ⊕ | ⊗ | ⊕ |
| McKnight et al., 2023 | US | ⊗ | ⊕ | ⊗ | ⊖ |
| McMahan et al., 2020 | US | ⊗ | ⊕ | ⊗ | ⊕ |
| Park et al., 2018 | US | ⊗ | ⊕ | ⊗ | ⊗ |
| Perez-Figueroa et al., 2022 | US | ⊗ | ⊗ | ⊗ | ⊕ |
| Pizzicato et al., 2020 | US | ⊗ | ⊕ | ⊗ | ⊕ |
| Riley et al., 2016 | US | ⊗ | ⊗ | ⊕ | ⊕ |
| Rosenthal. et al., 2020 | US | ⊗ | ⊗ | ○ | ○ |
| Shrestha et al., 2021 | US | ⊗ | ⊕ | ⊗ | ⊗ |
| Stephens et al., 2014 | US | ⊗ | ⊕ | ⊗ | ⊗ |
| Tesfaye-Rogoza et al., 2020 | US | ⊗ | ⊕ | ⊗ | ⊗ |
| Tomko et al., 2022 | US | ⊕ | ⊕ | ⊗ | ⊗ |
| Zhao et al., 2020 | US | ⊕ | ⊕ | ⊗ | ⊖ |

**Legend**

- ⊕ Low Risk of Bias
- ⊖ Unclear Risk of Bias
- ⊗ High Risk of Bias
- ○ Not Applicable

**Footnotes:**

<u>Abbreviations:</u> Person-Years (PY); North America (NA); United Kingdom (UK); United States (US)

Studies only presenting fatal overdose measures were excluded from the PY and definition risk of bias assessments.

* Participant recruitment conducted exclusively in the population (e.g., street recruitment) had low risk of bias. Studies that included participants recruited from venues (e.g., clinics, harm reduction programs) had high risk of bias.

§ Study sample sizes with ≥200 participants had low risk of bias and studies with sample sizes ≤199 participants had high risk of bias.

¶ Studies that reported PY directly had low risks of biases. Studies that did not report PY (i.e., PY had to be approximated) had high risks of biases.

† Definitions with low risk of bias were fully defined (e.g., [1] provides clear and concise references with [2] one or more limiting characteristics and [3] has an accompanying example) and overdose events without definitions had a high risk of bias. Studies that had partially defined (e.g., missing one or more definition components) overdose events that had uncertain risks of biases.

₪ Bowles et al., 2021; This reference contains data collected from multiple countries. Reference appears twice in this table, referring to Canada and Mexico respectively.

### **Appendix 4.** Results Tables

#### **Appendix 4.1:** Non-Fatal Overdose Sensitivity Analyses (2010 – 2023)

| **Location** | **N** | **Rate (95% CI)**[§, ₪, *] | **$I^2$ (%)** |
|---|---|---|---|
| **Overall** | 46 | 30.46 (25.25 – 36.74)₪ | 98.00 |
| Provided PY | 6 | 17.78 (12.40 – 25.47) | – |
| Approximated PY | 40 | 32.94 (27.02 – 40.16)* | – |
| 3 Month Recall | 2 | 56.14 (38.22 – 82.45) | – |
| 6 Month Recall | 18 | 40.95 (28.67 – 58.50) | – |
| 12 Month Recall | 20 | 25.88 (21.51 – 31.14)* | – |
| **United States** | 28 | 32.88 (26.43 – 40.91)₪ | 96.60 |
| Population-Based | 6 | 40.18 (23.96 – 67.38) | – |
| Venue-Based | 7 | 27.09 (21.12 – 34.76) | – |
| Mixed | 15 | 33.20 (23.63 – 46.64) | |

**Footnotes:**

Abbreviations: Sample Size (N); Confidence-Interval (CI); Person-years (PY)

* Between group differences p-value ≤ 0.05
₪ Q-test for heterogeneity p-value ≤ 0.05
§ Per 100 PY

### **Appendix 4.2:** Non-Fatal and Fatal Overdose Rates Estimates Among PWID (N=58)

| Reference | Location | Data Collection Years | Study Design | N | PY | Non-Fatal Rate (95% CI)[§] | Fatal Rate (95% CI)[‡,*,§] |
|---|---|---|---|---|---|---|---|
| College-Frisbey et al., 2023 | Australia | 2017-2021 | Cross-Sectional | 965 | 965† | 52.02 (47.66 — 56.78) | |
| Conway et al., 2021 | Australia | 2018-2019 | Cross-Sectional | 776 | 776† | 10.18 (8.17—12.69) | |
| Geddes et al., 2021 | Australia | 2019 | Cross-Sectional | 2609 | 2609† | 16.79 (15.29—18.44) | |
| Hill et al., 2022b | Australia | 2008-2019 | Longitudinal | 1209 | 6913 | | 0.51 (0.36—0.71) |
| Kimber et al., 2019 | Australia | 1999-2010 | Longitudinal | 215 | 2083 | | 0.24 (0.10—0.58) |
| Price et al., 2023 | Australia | 2012-2022 | Cross-Sectional | 5657 | 5657† | 17.96 (16.89—19.10) | |
| VanDenBoom et al., 2021 | Australia | 2018-2019 | Cross-Sectional | 658 | 658† | 9.42 (7.35—12.09) | |
| Bowles et al., 2021₪ | Canada | 2014-2018 | Cross-Sectional | 1328 | 664† | 22.59 (19.25—26.51) | |
| Hadland et al., 2014 | Canada | 2005-2012 | Longitudinal | 414 | 763 | 9.04 (7.14—11.45) | |
| Kennedy et al., 2019a | Canada | 2006-2017 | Longitudinal | 811 | 4872 | | 0.39 (0.25—0.61) |
| Kenny et al., 2022 | Canada | 2019 | Cross-Sectional | 243 | 122† | 97.54 (81.50—116.74) | |
| Martel-Laferriere et al., 2022 | Canada | 2017-2020 | Longitudinal | 62 | 24 | | 4.17 (0.59—29.58) |
| McCrae et al., 2020 | Canada | 2018 | Cross-Sectional | 793 | 397† | 32.24 (27.11—38.34) | |
| Roy et al., 2016 | Canada | 2012-2015 | Longitudinal | 122 | 31† | 48.39 (29.17—80.26) | |
| Rudzinski et al., 2021 | Canada | 2018-2019 | Cross-Sectional | 15 | 8† | 37.50 (12.09—116.27) | |
| Scheim et al., 2021 | Canada | 2018-2020 | Cross-Sectional | 699 | 350† | 77.14 (68.47—86.92) | |
| Selfridge et al., 2021 | Canada | 2014-2019 | Longitudinal | 232 | 377 | | 4.24 (2.60—6.93) |
| Shaw et al., 2015 | Canada | 2013 | Cross-Sectional | 272 | 272† | 29.04 (23.30—36.21) | |
| Tarasuk et al., 2020 | Canada | 2017-2019 | Cross-Sectional | 1652 | 826† | 45.28 (40.91—50.11) | |
| Vallance et al., 2018 | Canada | 2008-2015 | Cross-Sectional | 548 | 548† | 18.61 (15.33—22.60) | |
| Wallace et al., 2019 | Canada | 2016 | Cross-Sectional | 187 | 94† | 59.57 (45.85—77.41) | |
| Bowles et al., 2021₪ | Mexico | 2014-2018 | Cross-Sectional | 666 | 333† | 9.01 (6.30—12.89) | |
| West et al., 2020 | Mexico | 2011-2018 | Longitudinal | 658 | 3044 | | 0.46 (0.27—0.78) |
| Bailey et al., 2022 | Mexico & the US | 2020-2021 | Cross-Sectional | 612 | 306† | 31.70 (25.98—38.68) | |
| Byrne et al., 2022 | UK | 2017-2020 | Longitudinal | 231 | 1553 | | 1.87 (1.30—2.69) |
| McAuley et al., 2023 | UK | 2019-2020 | Cross-Sectional | 1632 | 1632† | 20.89 (18.79—23.23) | |
| McDonald et al., 2021 | UK | 2009-2018 | Longitudinal | 35065 | 236914 | | 0.80 (0.77—0.84) |
| Spring et al., 2022 | UK | 2019 | Cross-Sectional | 2139 | 2139† | 19.21 (17.44—21.16) | |
| Abadie et al., 2022 | US | 2015-2019 | Longitudinal | 66 | 132 | 17.42 (11.58—26.22) | |
| Ahmad et al., 2021 | US | 2018 | Cross-Sectional | 420 | 210† | 85.24 (73.62—98.69) | |
| Armenta et al., 2015 | US | 2012-2015 | Longitudinal | 574 | 676 | 15.09 (12.43—18.32) | |
| Bluthenthal et al., 2020 | US | 2016-2017 | Cross-Sectional | 814 | 407† | 55.53 (48.74—63.26) | |
| Buresh et al., 2017 | US | 2005-2013 | Longitudinal | 1310 | 2225 | 30.43 (28.22—32.81) | |
| Calvo et al., 2017 | US | 2014-2015 | Cross-Sectional | 107 | 107† | 28.04 (19.60—40.10) | |
| Carlson et al., 2020 | US | 2017-2018 | Cross-Sectional | 225 | 113† | 70.80 (56.86—88.14) | |
| Chandra et al., 2021 | US | 2018-2019 | Cross-Sectional | 104 | 104† | 19.23 (12.41—29.81) | |
| Davis et al., 2017 | US | 1996-2011 | Longitudinal | 2007 | 5339 | | 1.09 (0.84—1.41) |
| Glick et al., 2021 | US | 2017-2019 | Cross-Sectional | 720 | 720† | 46.53 (41.80—51.79) | |
| Hayes et al., 2022 | US | 2018-2021 | Cross-Sectional | 1028 | 1028† | 28.31 (25.23—31.75) | |
| Heimer et al., 2015 | US | 2008-2012 | Cross-Sectional | 456 | 456† | 23.03 (19.02—27.88) | |

### **Appendix 4.2:** Non-Fatal and Fatal Overdose Rates Estimates Among PWID (N=58)

| **Reference** | **Location** | **Data Collection Years** | **Study Design** | **N** | **PY** | **Non-Fatal Rate (95% CI)**[§] | **Fatal Rate (95% CI)**[‡,*,§] |
|---|---|---|---|---|---|---|---|
| Huang et al., 2018 | US | 2004-2017 | Longitudinal | 76 | 76[†] | | 1.32 (0.19—9.34) |
| Hunter et al., 2018 | US | 2016 | Cross-Sectional | 283 | 283[†] | 32.51 (26.50—39.88) | |
| Jones et al., 2022 | US | 2016-2019 | Cross-Sectional | 347 | 174[†] | 63.22 (52.44—76.21) | |
| Kinnard et al., 2021 | US | 2017-2018 | Cross-Sectional | 534 | 267[†] | 31.84 (25.74—39.38) | |
| Lambdin et al., 2022 | US | 2018-2020 | Longitudinal | 493 | 493[†] | | 2.23 (1.24—4.03) |
| Lin et al., 2023 | US | 2018-2021 | Cross-Sectional | 258 | 129[†] | 72.09 (58.83—88.34) | |
| Marcus et al., 2022 | US | 2018 | Cross-Sectional | 10197 | 10197[†] | 28.22 (27.21—29.27) | |
| McKnight et al., 2023 | US | 2021-2022 | Cross-Sectional | 313 | 157[†] | 45.86 (36.40—57.78) | |
| McMahan et al., 2020 | US | 2019 | Cross-Sectional | 583 | 583[†] | 24.87 (21.14—29.27) | |
| Park et al., 2018 | US | 2016 | Cross-Sectional | 203 | 203[†] | 31.03 (24.24—39.73) | |
| Perez-Figueroa et al., 2022 | US | 2019 | Cross-Sectional | 62 | 16[†] | 68.75 (38.07—124.14) | |
| Pizzicato et al., 2020 | US | 2017-2018 | Cross-Sectional | 299 | 299[†] | 47.16 (39.98—55.62) | |
| Riley et al., 2016 | US | 2012-2014 | Longitudinal | 173 | 140 | 19.29 (13.23—28.12) | |
| Rosenthal. et al., 2020 | US | 2017-2018 | Longitudinal | 100 | 46 | 23.91 (13.24—43.18) | 4.35 (1.09—17.38) |
| Shrestha et al., 2021 | US | 2017-2019 | Cross-Sectional | 279 | 279[†] | 47.31 (39.89—56.11) | |
| Stephens et al., 2014 | US | 2008-2010 | Cross-Sectional | 283 | 142[†] | 5.63 (2.82—11.27) | |
| Tesfaye-Rogoza et al., 2020 | US | 2017 | Cross-Sectional | 332 | 332[†] | 18.67 (14.56—23.95) | |
| Tomko et al., 2022 | US | 2018-2019 | Cross-Sectional | 238 | 119[†] | 74.79 (60.76—92.06) | |
| Zhao et al., 2020 | US | 2011-2013 | Cross-Sectional | 777 | 389[†] | 14.65 (11.30—19.00) | |

**Footnotes:**

Abbreviations: Sample Size (N); Person-Years (PY); Confidence Interval (CI)

† PY computed using the survey instrument's recall period.
‡ Fatal overdose measured over follow-up period.
* Excluding unconfirmed deaths.
§ Rates per 100 PY
₪ Bowles et al., 2021; This reference contains data collected from multiple countries. Reference appears twice in this table, referring to Canada and Mexico respectively.

### **Appendix 4.3:** Non-Analyzed Studies with Overlapping Data Summary Table (N=85)

| Reference | Location | Source of Overlapping Data Included in Analyses | Data Collection Years | N | PY | Non-Fatal Events | Fatal Events |
|---|---|---|---|---|---|---|---|
| Cunningham et al., 2018 | Australia | Martel-Laferriere et al., 2022; Selfridge et al., 2021 | 2016 | 103 | 71 | | 1 |
| Geddes et al., 2018 | Australia | Geddes et al., 2021 | 2014 | 838 | 838[†] | 211 | |
| Grebely et al., 2017 | Australia | Martel-Laferriere et al., 2022; Selfridge et al., 2021 | 2016 | 103 | 48 | | 1 |
| Grebely et al., 2018 | Australia | Martel-Laferriere et al., 2022; Selfridge et al., 2021 | 2016 | 102 | 6[†] | | 1 |
| Horyniak et al., 2013a | Australia | Price et al., 2023 | 2010-2011 | 991 | 496[†] | 53 | |
| Horyniak et al., 2013b | Australia | Colledge-Frisby et al., 2023 | 2008-2010 | 688 | 344[†] | 68 | |
| Iversen et al., 2017 | Australia | Geddes et al., 2021 | 2014 | 1488 | 1488[†] | 309 | |
| Nambiar et al., 2015 | Australia | Hill et al., 2022b; Colledge-Frisby et al., 2023 | 2008-2012 | 665 | 333; 2277[†] | 10 | 12 |
| Palmer et al., 2021 | Australia | Colledge-Frisby et al., 2023 | 2017-2019 | 720 | 720[†] | 59 | |
| Reddel et al., 2014 | Australia | Price et al., 2023 | 2011 | 200 | 200[†] | 13 | |
| Sutherland et al., 2021 | Australia | Price et al., 2023 | 2011-2018 | 1766 | 1766[†] | 297 | |
| Caudarella et al., 2016 | Canada | Bowles et al., 2021[¶]; Kennedy et al., 2019a | 1996-2011 | 2317 | 14996; 1499 | 1795 | 134 |
| Cheng et al., 2016 | Canada | Hadland et al., 2014 | 2008-2012 | 253 | 466 | 31 | |
| Escudero et al., 2016 | Canada | Bowles et al., 2021₪ | 2006-2013 | 1760 | 7535 | 649 | |
| Fairbairn et al., 2016 | Canada | Bowles et al., 2021₪ | 2005-2012 | 1114 | 4416 | 79 | |
| Gaddis et al., 2017 | Canada | Bowles et al., 2021₪ | 2010-2012 | 1316 | 658[†] | 49 | |
| Gower et al., 2022 | Canada | Scheim et al., 2021 | 2016 | 200 | 100[†] | 16 | |
| Hayashi et al., 2014 | Canada | Kennedy et al., 2019a | 1996-2011 | 2279 | 13597 | | 110 |
| Hayashi et al., 2016 | Canada | Kennedy et al., 2019a | 1996-2011 | 2317 | 14905 | | 120 |
| Hayashi et al., 2018 | Canada | Bowles et al., 2021₪ | 2016 | 669 | 335[†] | 85 | |
| Hayashi et al., 2021 | Canada | Bowles et al., 2021₪ | 2016-2017 | 590 | 295[†] | 127 | |
| Hyshka et al., 2015 | Canada | | | 279 | 140[†] | 62 | |
| Ickowicz et al., 2017 | Canada | Bowles et al., 2021₪ | 2005-2014 | 626 | 313[†] | 40 | |
| Ickowicz et al., 2020 | Canada | Bowles et al., 2021₪ | 2003-2017 | 793 | 397[†] | 86 | |
| Ickowicz et al., 2022 | Canada | Bowles et al., 2021₪ | 2017-2018 | 730 | 365[†] | 167 | |
| Kennedy et al., 2019b | Canada | Bowles et al., 2021₪ | 2005-2016 | 1336 | 6254 | 136 | |
| Kennedy et al., 2020 | Canada | Bowles et al., 2021₪ | 2017-2018 | 318 | 159[†] | 88 | |
| Kennedy et al., 2022 | Canada | Bowles et al., 2021₪ | 2015-2018 | 745 | 373[†] | 79 | |
| Lake et al., 2015a | Canada | Bowles et al., 2021₪ | 2005-2014 | 1660 | 5343 | 670 | |
| Lake et al., 2015b | Canada | Bowles et al., 2021₪ | 2005-2013 | 1697 | 4736 | 570 | |
| Lake et al., 2015c | Canada | Bowles et al., 2021₪ | 2005-2013 | 1614 | 4724 | 558 | |
| Mitra et al., 2022 | Canada | Schiem et al., 2021 | 2018-2020 | 701 | 351[†] | 270 | |
| Norton et al., 2022 | Canada | Bowles et al., 2021₪ | 2016-2018 | 1070 | 535[†] | 205 | |

### **Appendix 4.3:** Non-Analyzed Studies with Overlapping Data Summary Table (N=85)

| Reference | Location | Source of Overlapping Data Included in Analyses | Data Collection Years | N | PY | Non-Fatal Events | Fatal Events |
|---|---|---|---|---|---|---|---|
| Pedersen et al., 2016 | Canada | Bowles et al., 2021₪ | 2005-2014 | 1119 | 560† | 91 | |
| Prangnell et al., 2016 | Canada | Bowles et al., 2021₪ | 2005-2013 | 1142 | 571† | 72 | |
| Prangnell et al., 2019 | Canada | Bowles et al., 2021₪ | 2012-2016 | 327 | 164† | 47 | |
| Rammohan et al., 2022 | Canada | Schiem et al., 2021 | 2018-2019 | 604 | 302† | 224 | |
| Selfridge et al., 2019 | Canada | Selfridge et al., 2021 | 2014-2017 | 132 | 171 | | 9 |
| Tarasuk et al., 2021 | Canada | Tarasuk et al., 2020 | 2017-2019 | 997 | 499† | 185 | |
| Tucker et al., 2015 | Canada | Bowles et al., 2021₪ | 2005-2013 | 1911 | 956† | 133 | |
| Tucker et al., 2016 | Canada | Bowles et al., 2021₪ | 1996-2013 | 2806 | 1403† | 377 | |
| Collins et al., 2016∞ | Mexico | Bowles et al., 2021₪ | 2011-2014 | 735 | 368† | 74 | |
| Horyniak et al., 2018 | Mexico | Bowles et al., 2021₪ | 2011-2012 | 621 | 1921 | 59 | |
| Meacham et al., 2015§ | Mexico | West et al., 2020 | 2011-2012 | 735 | 368† | 74 | |
| Meacham et al., 2016 | Mexico | Armenta et al., 2015 | 2011-2014 | 1311 | 656† | 119 | |
| Meacham et al., 2018 | Mexico | Bowles et al., 2021₪ | 2011-2012 | 735 | 368† | 74 | |
| Rafful et al., 2015a | Mexico | Bowles et al., 2021₪ | 2013 | 366 | 183† | 29 | |
| Rafful et al., 2015b | Mexico | Bowles et al., 2021₪ | 2011 | 557 | 557 | 51 | |
| Rafful et al., 2017 | Mexico | Bowles et al., 2021₪ | 2011-2017 | 670 | 335† | 64 | |
| Rivera Saldana et al., 2021 | Mexico | Bowles et al., 2021₪ | 2011-2013 | 661 | 1029 | 115 | |
| West et al., 2018 | Mexico | West et al., 2020 | 2011-2017 | 863 | 3132 | | 35 |
| Byrne et al., 2020 | UK | Byrne et al., 2022 | 2018-2020 | 129 | 30 | | 1 |
| Dunleavy et al., 2021 | UK | McAuley et al., 2023 | 2017-2018 | 1408 | 1408† | 260 | |
| Hope et al., 2016 | UK | Spring et al., 2022 | 2013 | 2047 | 2047† | 277 | |
| O'Halloran et al., 2017 | UK | Spring et al., 2022 | 2013-2014 | 3850 | 3850† | 591 | |
| Trayner et al., 2020 | UK | McAuley et al., 2023 | 2017-2018 | 1437 | 719† | 264 | |
| Trayner et al., 2021 | UK | McAuley et al., 2023 | 2017-2018 | 1421 | 1421† | 261 | |
| Yuan et al., 2022 | UK | Spring et al., 2022 | 2019 | 1972 | 1972† | 392 | |
| Allen et al., 2019 | US | Ahmad et al., 2021 | 2018 | 371 | 186† | 162 | |
| Al-Tayyib et al., 2017a | US | Marcus et al., 2022 | 2015 | 599 | 599† | 128 | |
| Al-Tayyib et al., 2017b | US | Marcus et al., 2022 | 2015 | 592 | 592† | 129 | |
| Arreola et al., 2014 | US | Zhao et al., 2020 | 2011-2013 | 696 | 348† | 46 | |
| Cedarbaum et al., 2014 | US | Glick et al., 2021 | 2013 | 389 | 389† | 94 | |
| Cedarbaum et al., 2016 | US | Glick et al., 2021 | 2013 | 389 | 389† | 95 | |
| Chiang et al., 2022 | US | Bluthenthal et al., 2020 | 2016-2018 | 601 | 301† | 85 | |
| Collins et al., 2016∞ | US | Armenta et al., 2015 | 2012-2014 | 291 | 146† | 23 | |
| Frost et al., 2017 | US | McMahn et al., 2020 | 2015 | 438 | 438† | 98 | |
| Glick et al., 2018 | US | Glick et al., 2021; Marcus et al., 2022 | 2009-2017 | 3844 | 3844† | 623 | |

### **Appendix 4.3:** Non-Analyzed Studies with Overlapping Data Summary Table (N=85)

| Reference | Location | Source of Overlapping Data Included in Analyses | Data Collection Years | N | PY | Non-Fatal Events | Fatal Events |
|---|---|---|---|---|---|---|---|
| Heimer et al., 2014 | US | Heimer et al., 2015 | 2008-2011 | 454 | 454[†] | 18 | |
| Hill et al., 2022a | US | Rosenthal et al., 2020 | 2017 | 49 | 49[†] | 8 | |
| Jarlais et al., 2023 | US | McKnight et al., 2023 | 2021-2022 | 275 | 138[†] | 59 | |
| Lipira et al., 2021 | US | Marcus et al., 2022 | 2018-2019 | 610 | 610[†] | 156 | |
| Mazhnaya et al., 2020 | US | Ahmad et al., 2021 | 2018 | 311 | 156[†] | 161 | |
| Meacham et al., 2015[§] | US | Armenta et al., 2015 | 2012-2014 | 576 | 288[†] | 45 | |
| Oh et al., 2020 | US | Marcus et al., 2022 | 2018 | 458 | 458[†] | 233 | |
| O'Rourke et al., 2019 | US | Ahmad et al., 2021 | 2018 | 373 | 187[†] | 163 | |
| Reid et al., 2023 | US | Glick et al., 2021; Marcus et al., 2022 | 2018 | 550 | 550[†] | 143 | |
| Rivera et al., 2022 | US | Marcus et al., 2022 | 2018 | 503 | 503[†] | 125 | |
| Robinson et al., 2017 | US | Marcus et al., 2022 | 2012 | 1708 | 1708 | 232 | |
| Rosenthal et al., 2019 | US | Rosenthal et al., 2020 | | 100 | 100 | 11 | 4 |
| Rouhani et al., 2021 | US | Ahmad et al., 2021 | 2018 | 420 | 210[†] | 179 | |
| Rushmore et al., 2023 | US | Marcus et al., 2022 | 2018 | 4420 | 4420[†] | 1344 | |
| Simpson et al., 2021 | US | Bluthenthal et al., 2021 | 2016-2018 | 531 | 266[†] | 84 | |
| Tsui et al., 2018 | US | Glick et al., 2021; Marcus et al., 2022 | 2015 | 486 | 486[†] | 106 | |
| Wagner et al., 2014 | US | Bowles et al., 2021₪ | 2012-2013 | 480 | 240[†] | 39 | |
| Wagner et al., 2015 | US | Bowles et al., 2021₪ | 2012-2014 | 573 | 287[†] | 45 | |
| Nahass et al., 2021 | | | | 30 | 30[†] | | 1 |

**Footnotes:**

Abbreviations: Sample Size (N); Person-Years (PY)

All data reported here were excluded from the meta-analysis.

[†] PY computed using the survey instrument's recall period.

[∞] Collins et al., 2016; This reference contains data collected from multiple countries. This reference appears multiple times in this table as this meta-analysis analyzes data on country level differences.

[§] Meacham et al., 2015; This reference contains data collected from multiple countries. This reference appears multiple times in this table as this meta-analysis analyzes data on country level differences.

[₪] Bowles et al., 2021; This reference contains data collected from multiple countries. This reference appears multiple times in this table as this meta-analysis analyzes data on country level differences.

**Appendix 5.** Overall Analyses Forest Plots

| Reference | Rate | 95% CI | Date Range |
|---|---|---|---|
| Buresh et al. 2017 | 30.43 | [28.22; 32.81] | 2005-2013 |
| Hadland et al. 2014 | 9.04 | [ 7.14; 11.45] | 2005-2012 |
| Stephens et al. 2014 | 5.63 | [ 2.82; 11.27] | 2008-2010 |
| Heimer et al. 2015 | 23.03 | [19.02; 27.88] | 2008-2012 |
| Vallance et al. 2018 | 18.61 | [15.33; 22.60] | 2008-2015 |
| Zhao et al. 2020 | 14.65 | [11.30; 19.00] | 2011-2013 |
| Riley et al. 2016 | 19.29 | [13.23; 28.12] | 2012-2014 |
| Shaw et al. 2015 | 29.04 | [23.30; 36.21] | 2013 |
| Armenta et al. 2015 | 15.09 | [12.43; 18.32] | 2012-2015 |
| Roy et al. 2016 | 48.39 | [29.17; 80.26] | 2012-2015 |
| Calvo et al. 2017 | 28.04 | [19.60; 40.10] | 2014-2015 |
| Bowles et al. 2021[pt1] | 9.01 | [ 6.30; 12.89] | 2014-2018 |
| Bowles et al. 2021[pt2] | 22.59 | [19.25; 26.51] | 2014-2018 |
| Hunter et al. 2018 | 32.51 | [26.50; 39.88] | 2016 |
| Park et al. 2018 | 31.03 | [24.24; 39.73] | 2016 |
| Wallace et al. 2019 | 59.57 | [45.85; 77.41] | 2016 |
| Price et al. 2023 | 17.96 | [16.89; 19.10] | 2012-2022 |
| Abadie et al. 2022 | 17.42 | [11.58; 26.22] | 2015-2019 |
| Bluthenthal et al. 2020 | 55.53 | [48.74; 63.26] | 2016-2017 |
| Tesfaye-Rogoza et al. 2020 | 18.67 | [14.56; 23.95] | 2017 |
| Jones et al. 2022 | 63.22 | [52.44; 76.21] | 2016-2019 |
| Carlson et al. 2020 | 70.80 | [56.86; 88.14] | 2017-2018 |
| Glick et al. 2021 | 46.53 | [41.80; 51.79] | 2017-2019 |
| Kinnard et al. 2021 | 31.84 | [25.74; 39.38] | 2017-2018 |
| Pizzicato et al. 2020 | 47.16 | [39.98; 55.62] | 2017-2018 |
| Rosenthal et al. 2020 | 23.91 | [13.24; 43.18] | 2017-2018 |
| Shrestha et al. 2021 | 47.31 | [39.89; 56.11] | 2017-2019 |
| Tarasuk et al. 2020 | 45.28 | [40.91; 50.11] | 2017-2019 |
| Ahmad et al. 2021 | 85.24 | [73.62; 98.69] | 2018 |
| Marcus et al. 2022 | 28.22 | [27.21; 29.27] | 2018 |
| Colledge-Frisby et al. 2023 | 52.02 | [47.66; 56.78] | 2017-2021 |
| Chandra et al. 2021 | 19.23 | [12.41; 29.81] | 2018-2019 |
| Conway et al. 2021 | 10.18 | [ 8.17; 12.69] | 2018-2019 |
| Rudzinski et al. 2021 | 37.50 | [12.09; 116.27] | 2018-2019 |
| Scheim et al. 2021 | 77.14 | [68.47; 86.92] | 2018-2020 |
| Tomko et al. 2022 | 74.79 | [60.76; 92.06] | 2018-2019 |
| Geddes et al. 2021 | 16.79 | [15.29; 18.44] | 2019 |
| Kenny et al. 2022 | 97.54 | [81.50; 116.74] | 2019 |
| McMahan et al. 2020 | 24.87 | [21.14; 29.27] | 2019 |
| Perez-Figueroa et al. 2022 | 68.75 | [38.07; 124.14] | 2019 |
| Spring et al. 2022 | 19.21 | [17.44; 21.16] | 2019 |
| Hayes et al. 2022 | 28.31 | [25.23; 31.75] | 2018-2021 |
| Lin et al. 2023 | 72.09 | [58.83; 88.34] | 2018-2021 |
| McAuley et al. 2023 | 20.89 | [18.79; 23.23] | 2019-2020 |
| Bailey et al. 2022 | 31.70 | [25.98; 38.68] | 2020-2021 |
| McKnight et al. 2023 | 45.86 | [36.40; 57.78] | 2021-2022 |
| **Random effects model** | **30.46** | **[25.25; 36.74]** | |

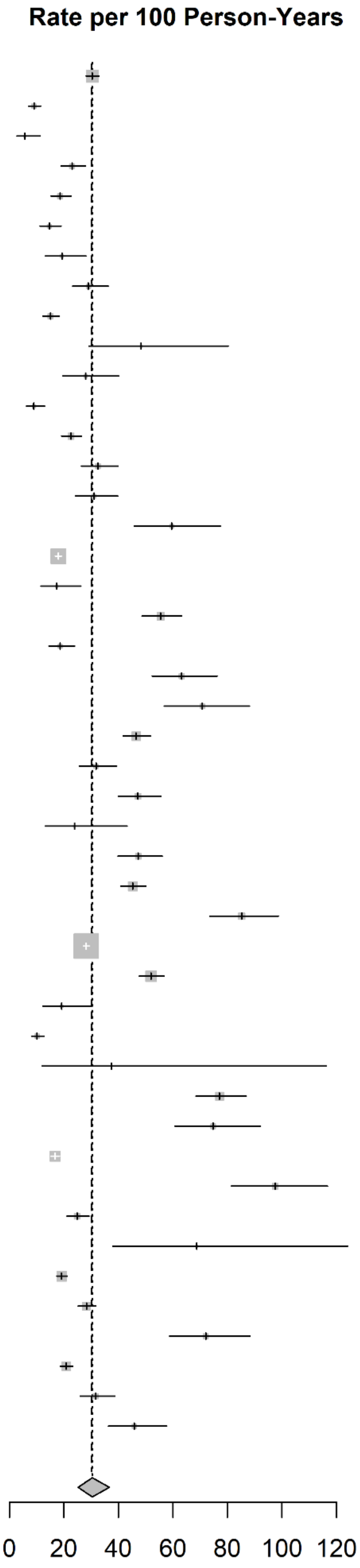

**Appendix 5.1:** Random-Effects Meta-Analysis of North American, United Kingdom, and Australian Non-Fatal Overdose Rates Per 100 PY

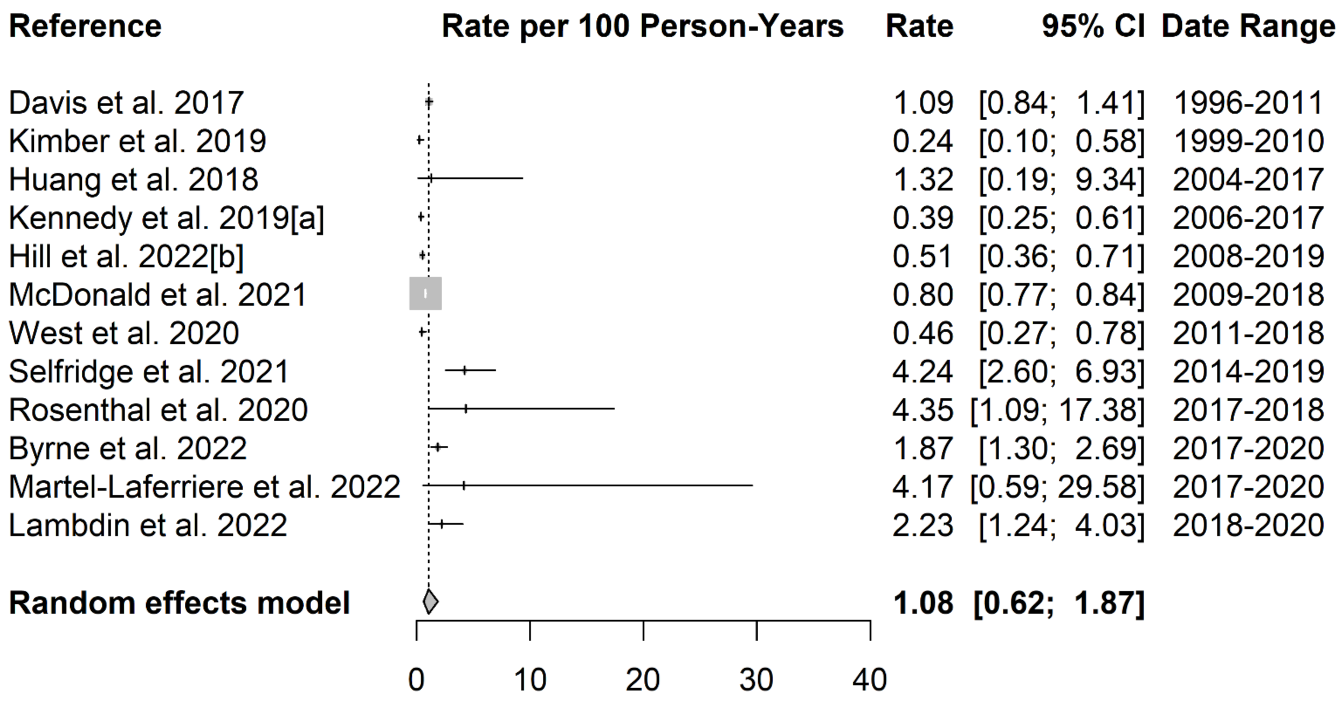

**Appendix 5.2:** Random-Effects Meta-Analysis of North American, United Kingdom, and Australian Fatal Overdose Rates Per 100 PY

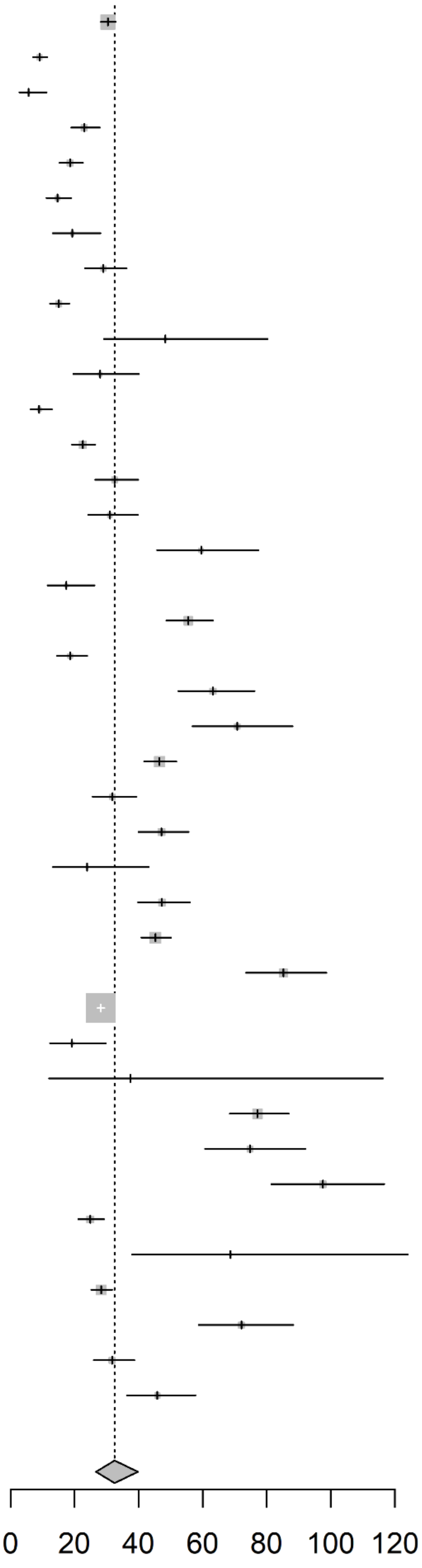

| Reference | Rate | 95% CI | Date Range |
|---|---|---|---|
| Buresh et al. 2017 | 30.43 | [28.22; 32.81] | 2005-2013 |
| Hadland et al. 2014 | 9.04 | [ 7.14; 11.45] | 2005-2012 |
| Stephens et al. 2014 | 5.63 | [ 2.82; 11.27] | 2008-2010 |
| Heimer et al. 2015 | 23.03 | [19.02; 27.88] | 2008-2012 |
| Vallance et al. 2018 | 18.61 | [15.33; 22.60] | 2008-2015 |
| Zhao et al. 2020 | 14.65 | [11.30; 19.00] | 2011-2013 |
| Riley et al. 2016 | 19.29 | [13.23; 28.12] | 2012-2014 |
| Shaw et al. 2015 | 29.04 | [23.30; 36.21] | 2013 |
| Armenta et al. 2015 | 15.09 | [12.43; 18.32] | 2012-2015 |
| Roy et al. 2016 | 48.39 | [29.17; 80.26] | 2012-2015 |
| Calvo et al. 2017 | 28.04 | [19.60; 40.10] | 2014-2015 |
| Bowles et al. 2021[pt1] | 9.01 | [ 6.30; 12.89] | 2014-2018 |
| Bowles et al. 2021[pt2] | 22.59 | [19.25; 26.51] | 2014-2018 |
| Hunter et al. 2018 | 32.51 | [26.50; 39.88] | 2016 |
| Park et al. 2018 | 31.03 | [24.24; 39.73] | 2016 |
| Wallace et al. 2019 | 59.57 | [45.85; 77.41] | 2016 |
| Abadie et al. 2022 | 17.42 | [11.58; 26.22] | 2015-2019 |
| Bluthenthal et al. 2020 | 55.53 | [48.74; 63.26] | 2016-2017 |
| Tesfaye-Rogoza et al. 2020 | 18.67 | [14.56; 23.95] | 2017 |
| Jones et al. 2022 | 63.22 | [52.44; 76.21] | 2016-2019 |
| Carlson et al. 2020 | 70.80 | [56.86; 88.14] | 2017-2018 |
| Glick et al. 2021 | 46.53 | [41.80; 51.79] | 2017-2019 |
| Kinnard et al. 2021 | 31.84 | [25.74; 39.38] | 2017-2018 |
| Pizzicato et al. 2020 | 47.16 | [39.98; 55.62] | 2017-2018 |
| Rosenthal et al. 2020 | 23.91 | [13.24; 43.18] | 2017-2018 |
| Shrestha et al. 2021 | 47.31 | [39.89; 56.11] | 2017-2019 |
| Tarasuk et al. 2020 | 45.28 | [40.91; 50.11] | 2017-2019 |
| Ahmad et al. 2021 | 85.24 | [73.62; 98.69] | 2018 |
| Marcus et al. 2022 | 28.22 | [27.21; 29.27] | 2018 |
| Chandra et al. 2021 | 19.23 | [12.41; 29.81] | 2018-2019 |
| Rudzinski et al. 2021 | 37.50 | [12.09; 116.27] | 2018-2019 |
| Scheim et al. 2021 | 77.14 | [68.47; 86.92] | 2018-2020 |
| Tomko et al. 2022 | 74.79 | [60.76; 92.06] | 2018-2019 |
| Kenny et al. 2022 | 97.54 | [81.50; 116.74] | 2019 |
| McMahan et al. 2020 | 24.87 | [21.14; 29.27] | 2019 |
| Perez-Figueroa et al. 2022 | 68.75 | [38.07; 124.14] | 2019 |
| Hayes et al. 2022 | 28.31 | [25.23; 31.75] | 2018-2021 |
| Lin et al. 2023 | 72.09 | [58.83; 88.34] | 2018-2021 |
| Bailey et al. 2022 | 31.70 | [25.98; 38.68] | 2020-2021 |
| McKnight et al. 2023 | 45.86 | [36.40; 57.78] | 2021-2022 |
| **Random effects model** | **32.54** | **[26.64; 39.74]** | |

(Plot column header: **Rate per 100 Person-Years**)

**Appendix 5.3:** Random-Effects Meta-Analysis of North American Non-Fatal Overdose Rates Per 100 PY

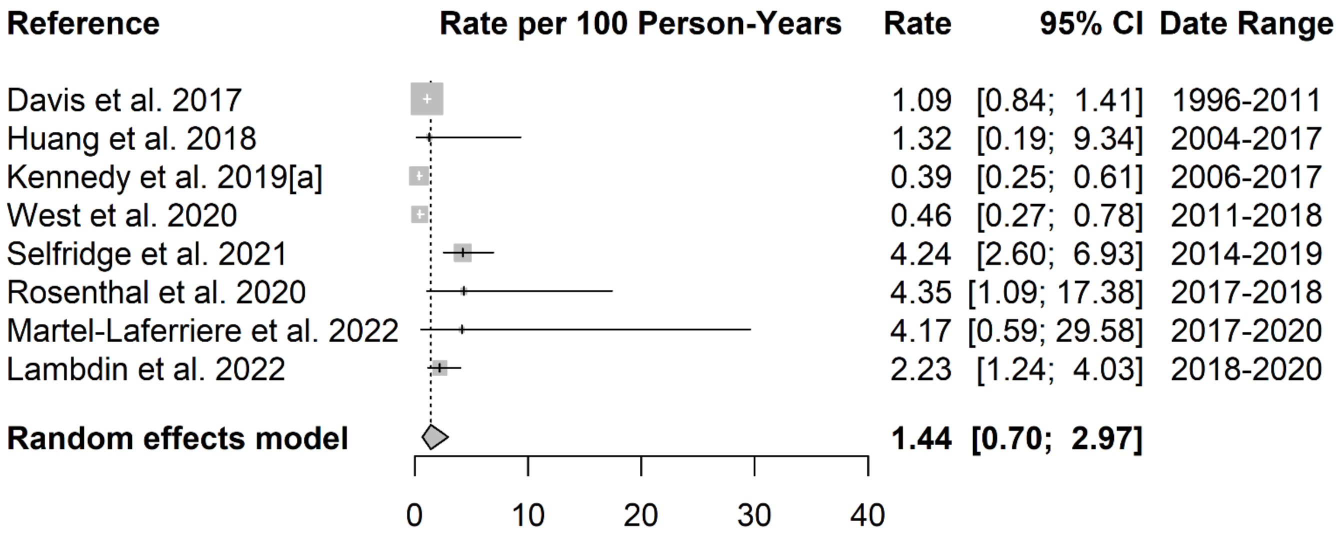

**Appendix 5.4:** Random-Effects Meta-Analysis of North American Fatal Overdose Rates Per 100 PY

**Appendix 6.** Post 2016 Analyses Forest Plots

| Reference | Rate | 95% CI | Date Range |
|---|---|---|---|
| Tesfaye-Rogoza et al. 2020 | 18.67 | [14.56; 23.95] | 2017 |
| Carlson et al. 2020 | 70.80 | [56.86; 88.14] | 2017-2018 |
| Glick et al. 2021 | 46.53 | [41.80; 51.79] | 2017-2019 |
| Kinnard et al. 2021 | 31.84 | [25.74; 39.38] | 2017-2018 |
| Pizzicato et al. 2020 | 47.16 | [39.98; 55.62] | 2017-2018 |
| Rosenthal et al. 2020 | 23.91 | [13.24; 43.18] | 2017-2018 |
| Shrestha et al. 2021 | 47.31 | [39.89; 56.11] | 2017-2019 |
| Tarasuk et al. 2020 | 45.28 | [40.91; 50.11] | 2017-2019 |
| Ahmad et al. 2021 | 85.24 | [73.62; 98.69] | 2018 |
| Marcus et al. 2022 | 28.22 | [27.21; 29.27] | 2018 |
| McCrae et al. 2020 | 32.24 | [27.11; 38.34] | 2018 |
| Colledge-Frisby et al. 2023 | 52.02 | [47.66; 56.78] | 2017-2021 |
| Chandra et al. 2021 | 19.23 | [12.41; 29.81] | 2018-2019 |
| Conway et al. 2021 | 10.18 | [ 8.17; 12.69] | 2018-2019 |
| Rudzinski et al. 2021 | 37.50 | [12.09; 116.27] | 2018-2019 |
| Scheim et al. 2021 | 77.14 | [68.47; 86.92] | 2018-2020 |
| Tomko et al. 2022 | 74.79 | [60.76; 92.06] | 2018-2019 |
| VanDenBoom et al. 2021 | 9.42 | [ 7.35; 12.09] | 2018-2019 |
| Geddes et al. 2021 | 16.79 | [15.29; 18.44] | 2019 |
| Kenny et al. 2022 | 97.54 | [81.50; 116.74] | 2019 |
| McMahan et al. 2020 | 24.87 | [21.14; 29.27] | 2019 |
| Perez-Figueroa et al. 2022 | 68.75 | [38.07; 124.14] | 2019 |
| Spring et al. 2022 | 19.21 | [17.44; 21.16] | 2019 |
| Hayes et al. 2022 | 28.31 | [25.23; 31.75] | 2018-2021 |
| Lin et al. 2023 | 72.09 | [58.83; 88.34] | 2018-2021 |
| McAuley et al. 2023 | 20.89 | [18.79; 23.23] | 2019-2020 |
| Bailey et al. 2022 | 31.70 | [25.98; 38.68] | 2020-2021 |
| McKnight et al. 2023 | 45.86 | [36.40; 57.78] | 2021-2022 |
| **Random effects model** | **35.51** | **[28.05; 44.96]** | |

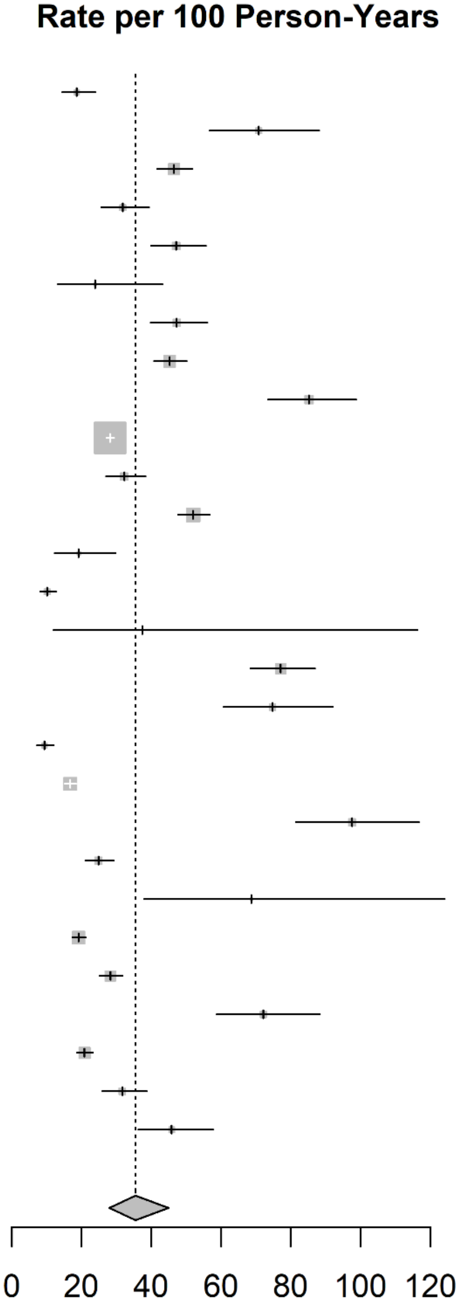

**Appendix 6.1:** Random-Effects Meta-Analysis of Post-2016 North American, United Kingdom, and Australian Non-Fatal Overdose Rates Per 100 PY

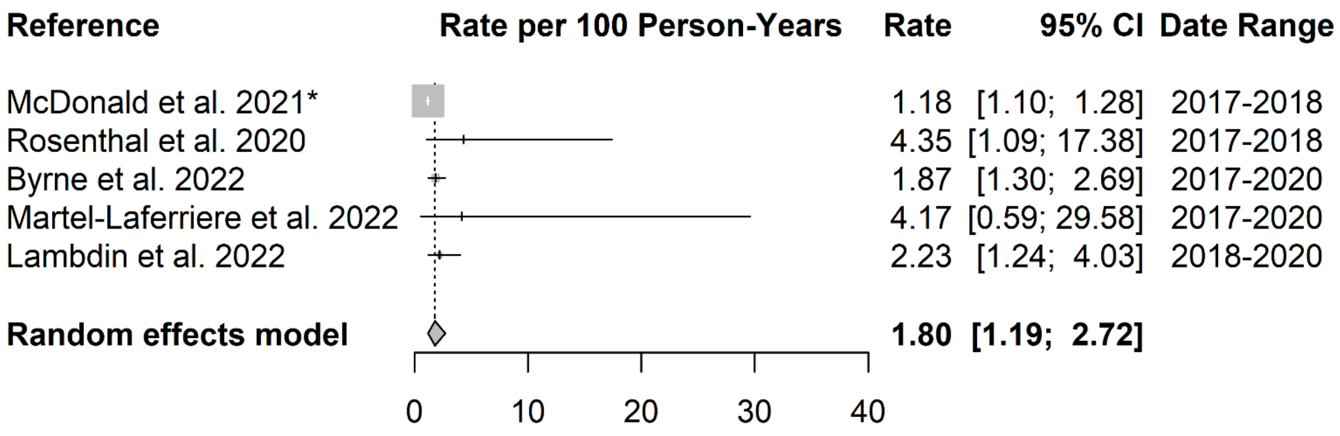

**Appendix 6.2:** Random-Effects Meta-Analysis of Post-2016 North American, United Kingdom, and Australian Fatal Overdose Rates Per 100 PY

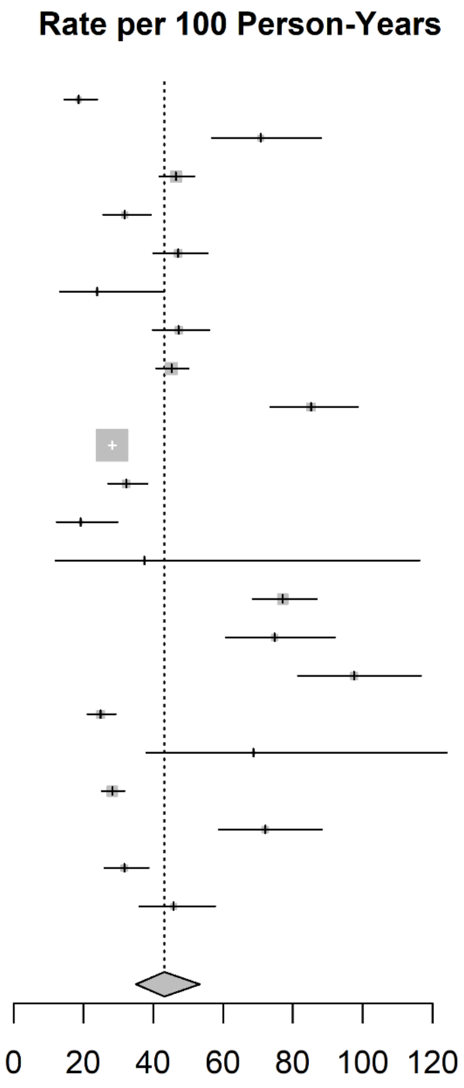

| Reference | Rate | 95% CI | Date Range |
|---|---|---|---|
| Tesfaye-Rogoza et al. 2020 | 18.67 | [14.56; 23.95] | 2017 |
| Carlson et al. 2020 | 70.80 | [56.86; 88.14] | 2017-2018 |
| Glick et al. 2021 | 46.53 | [41.80; 51.79] | 2017-2019 |
| Kinnard et al. 2021 | 31.84 | [25.74; 39.38] | 2017-2018 |
| Pizzicato et al. 2020 | 47.16 | [39.98; 55.62] | 2017-2018 |
| Rosenthal et al. 2020 | 23.91 | [13.24; 43.18] | 2017-2018 |
| Shrestha et al. 2021 | 47.31 | [39.89; 56.11] | 2017-2019 |
| Tarasuk et al. 2020 | 45.28 | [40.91; 50.11] | 2017-2019 |
| Ahmad et al. 2021 | 85.24 | [73.62; 98.69] | 2018 |
| Marcus et al. 2022 | 28.22 | [27.21; 29.27] | 2018 |
| McCrae et al. 2020 | 32.24 | [27.11; 38.34] | 2018 |
| Chandra et al. 2021 | 19.23 | [12.41; 29.81] | 2018-2019 |
| Rudzinski et al. 2021 | 37.50 | [12.09; 116.27] | 2018-2019 |
| Scheim et al. 2021 | 77.14 | [68.47; 86.92] | 2018-2020 |
| Tomko et al. 2022 | 74.79 | [60.76; 92.06] | 2018-2019 |
| Kenny et al. 2022 | 97.54 | [81.50; 116.74] | 2019 |
| McMahan et al. 2020 | 24.87 | [21.14; 29.27] | 2019 |
| Perez-Figueroa et al. 2022 | 68.75 | [38.07; 124.14] | 2019 |
| Hayes et al. 2022 | 28.31 | [25.23; 31.75] | 2018-2021 |
| Lin et al. 2023 | 72.09 | [58.83; 88.34] | 2018-2021 |
| Bailey et al. 2022 | 31.70 | [25.98; 38.68] | 2020-2021 |
| McKnight et al. 2023 | 45.86 | [36.40; 57.78] | 2021-2022 |
| **Random effects model** | **43.21** | **[35.05; 53.28]** | |

**Appendix 6.3:** Random-Effects Meta-Analysis of Post-2016 North American Non-Fatal Overdose Rates Per 100 PY

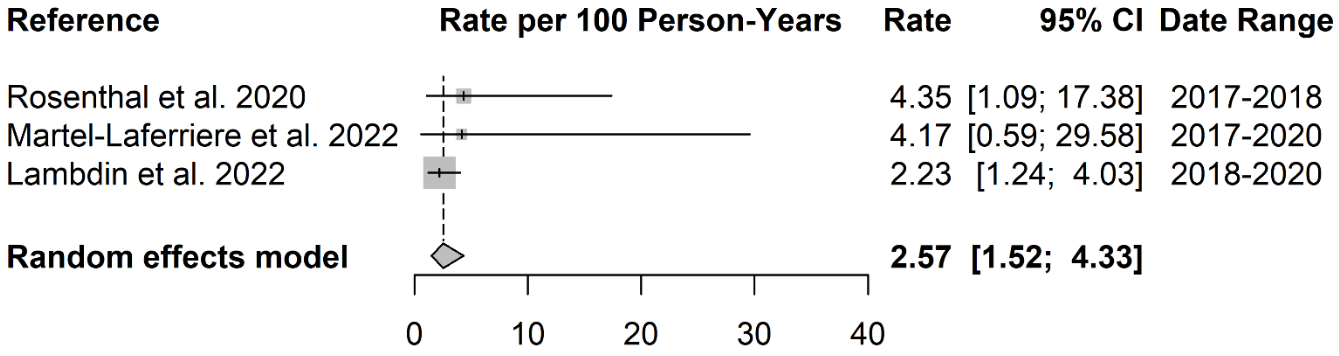

**Appendix 6.4:** Random-Effects Meta-Analysis of Post-2016 North American Fatal Overdose Rates Per 100 PY

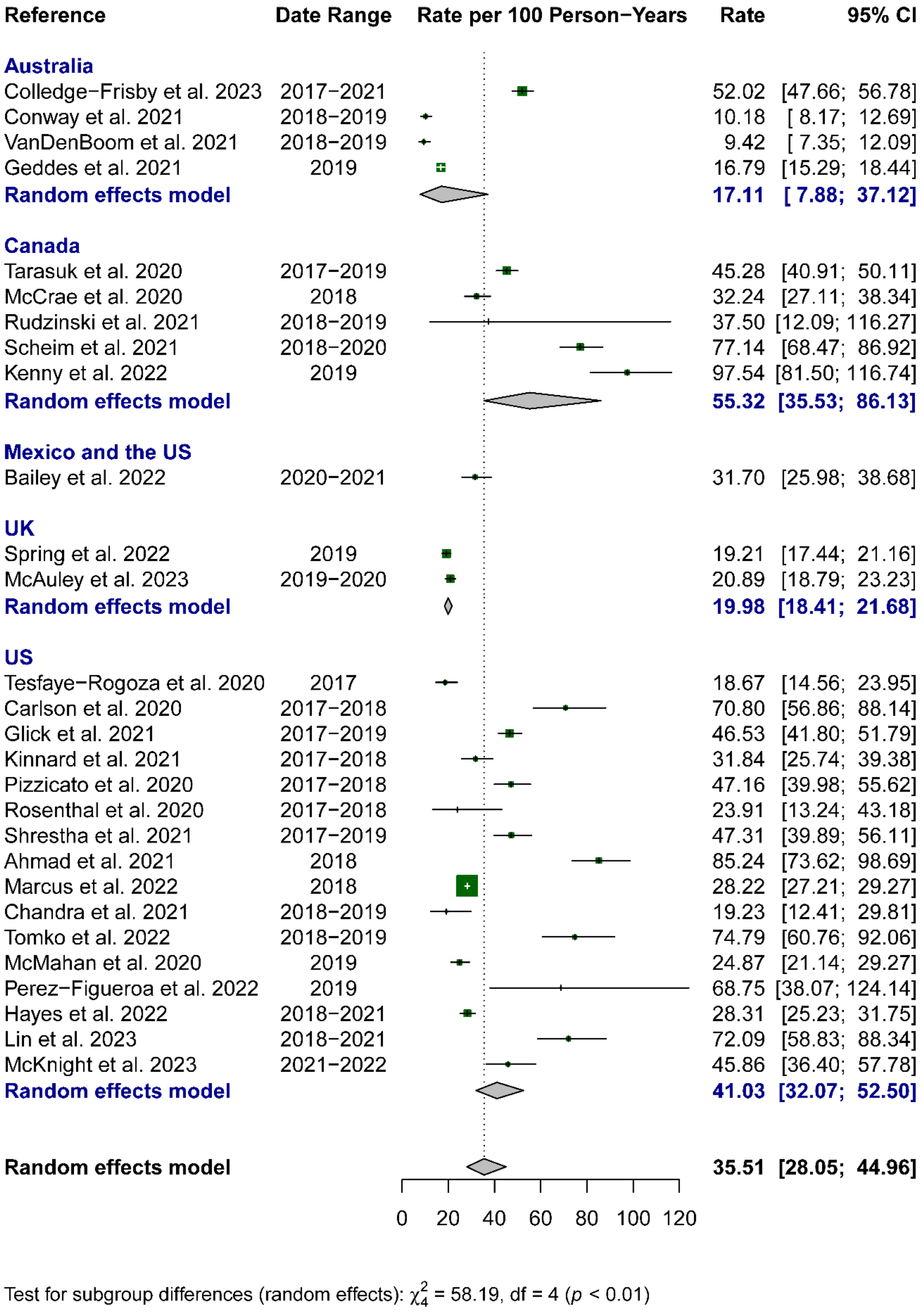

**Appendix 6.5:** Random-Effects Meta-Analysis of Post-2016 Country-Level Non-Fatal Overdose Rates Per 100 PY

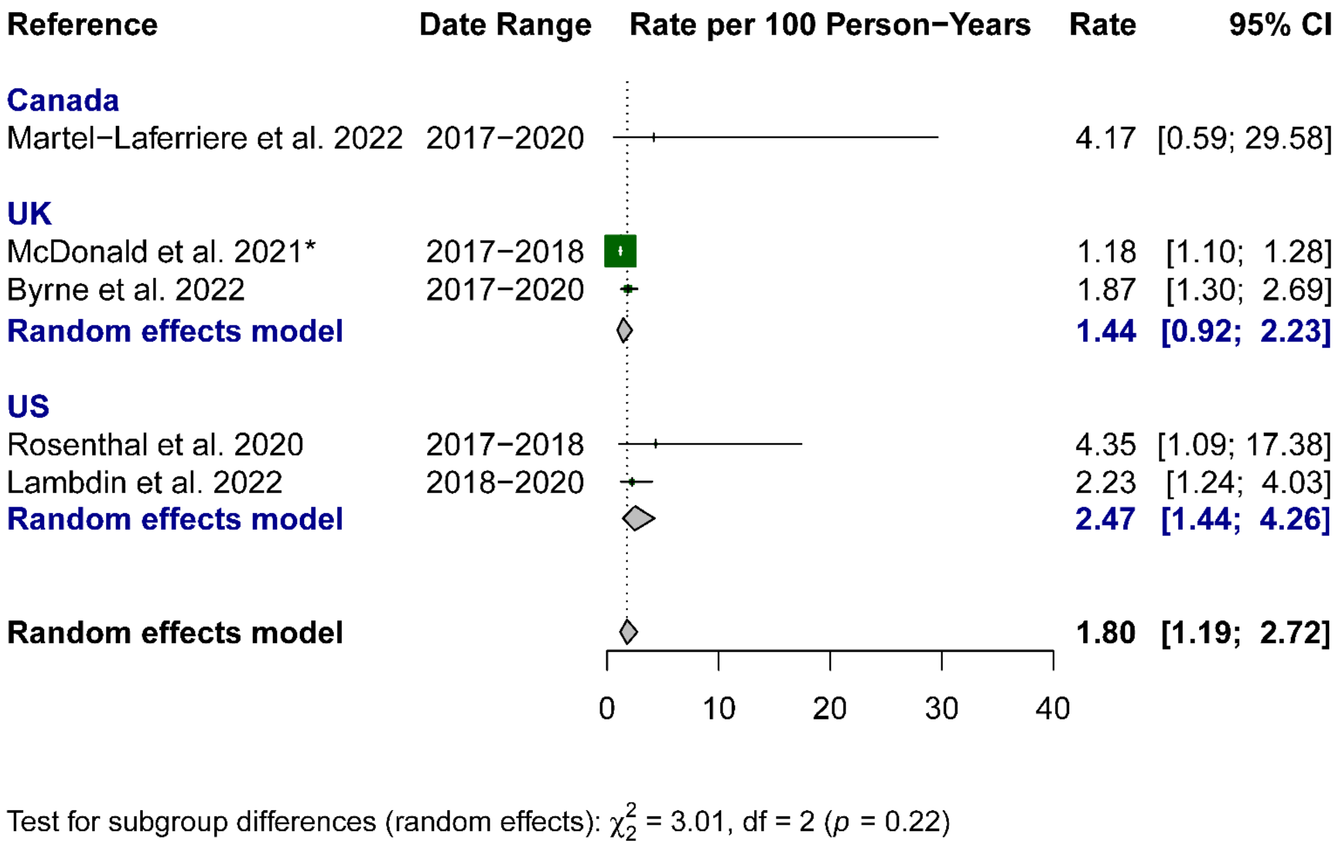

**Appendix 6.6:** Random-Effects Meta-Analysis of Post-2016 Country-Level Fatal Overdose Rates Per 100 PY

**Appendix 7.** Line Plots

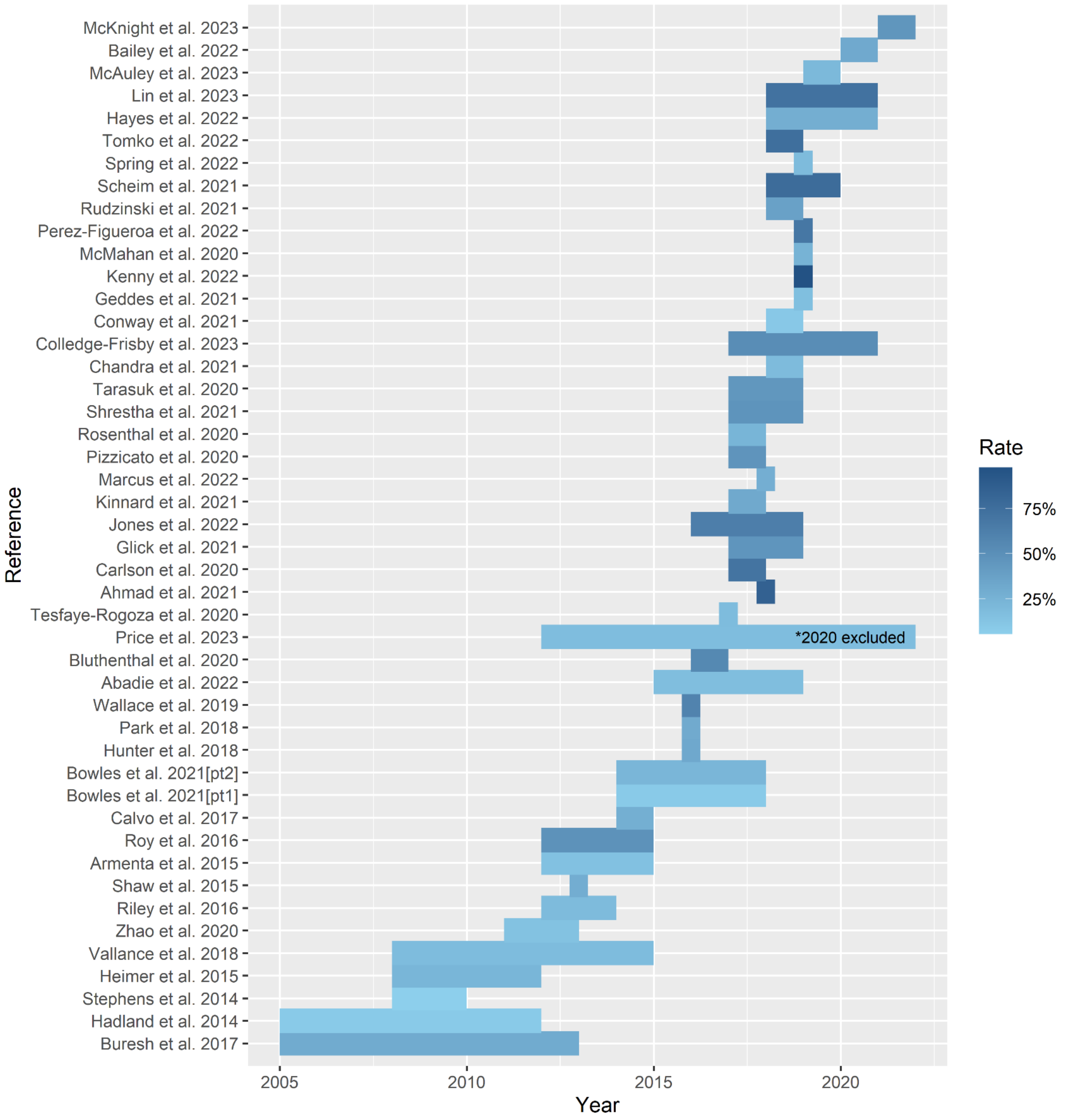

**Appendix 7.1:** Line Plot of North American, United Kingdom, and Australian Non-Fatal Overdose Rates Per 100 PY

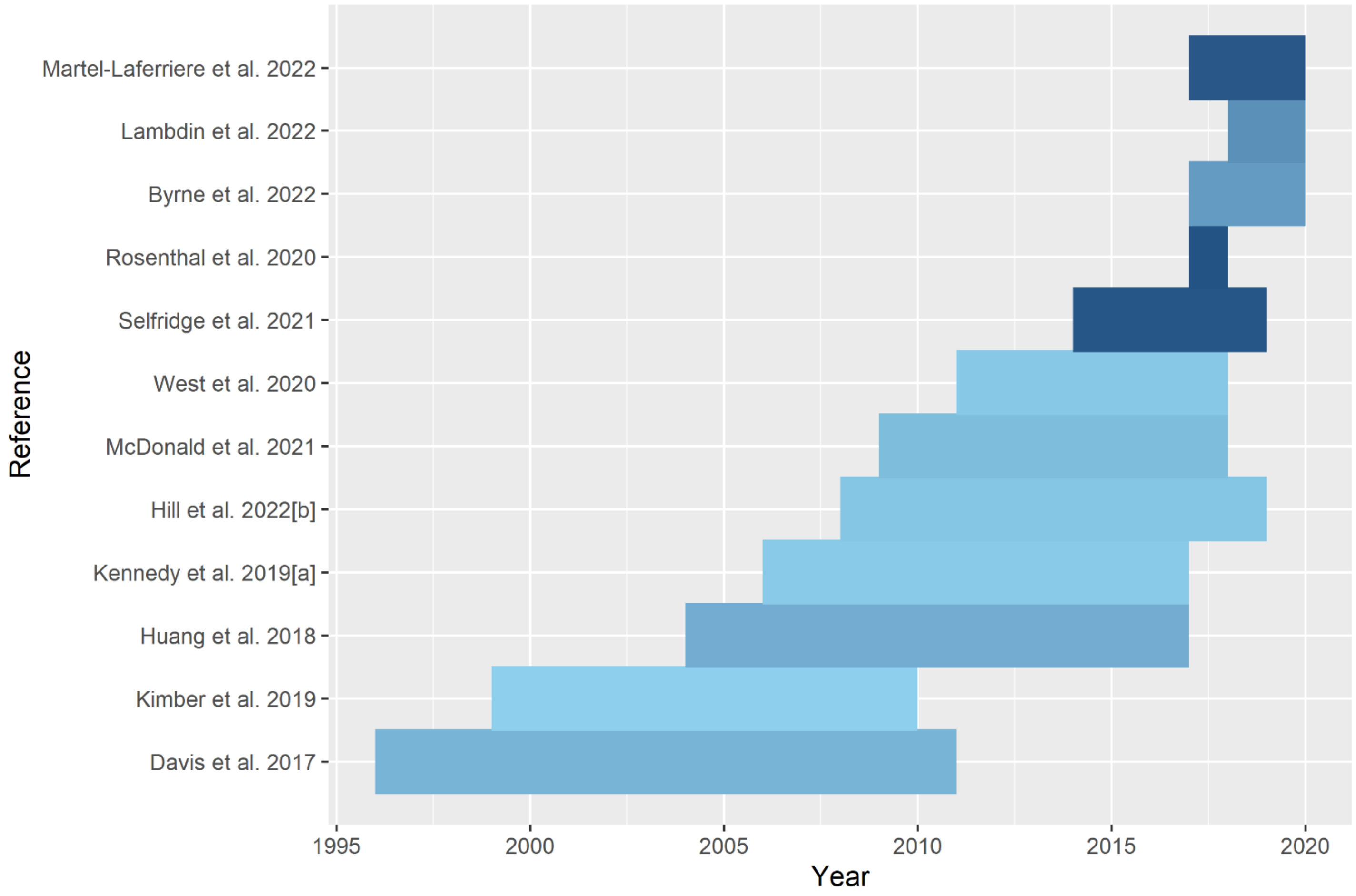

**Appendix 7.2:** Line Plot of North American, United Kingdom, and Australian Fatal Overdose Rates Per 100 PY
